# Genomic Signatures and Prediction of Clinical Severity in *Klebsiella pneumoniae* infections in a Multicenter Cohort

**DOI:** 10.64898/2026.02.02.26345332

**Authors:** Mohammad Malaikah, Rfeef Yousf Alyami, Jiayi Huang, Omniya Fallatah, Mathew Milner, Ge Zhou, Raneem Hirayban, Sara Iftikhar, Manuel Banzhaf, Yan Li, Abiola Senok, Sharif Matouq Hala, Nouf Batook, Dhuha Alsharif, Alhanouf Sultan AlShahrani, Abdulfattah Wasel Alamri, Sameera M AlJohani, Mai M Kaaki, Basaam Alwan, Mohammad Absar, Mohammad Elsir Al. Ali, Haitham S Sadah, Samer Zakri, Mohammad Bosaeed, Arnab Pain, Danesh Moradigaravand

## Abstract

*Klebsiella pneumoniae* is a major causative agent of hospital-acquired infections worldwide, contributing substantially to morbidity, mortality, and healthcare burden.. The emergence of strains that combine resistance to last-resort antimicrobials with hypervirulence has become a pressing public-health challenge. Despite extensive characterization of the genetic determinants of multidrug resistance and hypervirulence, the relationship between the genetic repertoire of *K. pneumoniae* and the clinical severity of infections remains inadequately understood.

**Methods:** We analyzed a nationwide large-scale collection of 1,306 *K. pneumoniae* complex strains retrieved over seven years from five centres across the Kingdom of Saudi Arabia. Using detailed and comprehensive patient-level clinical data, We employed a range of regression analyses, genome-wide association study (GWAS) methods, and machine-learning approaches to elucidate the clinical significance of ESBL/carbapenemase-producing (ESBL/CP), hypervirulent, and convergent ESBL(+)/CP(+) hypervirulent *K. pneumoniae* strains. We examined clinical severity outcomes including in-hospital all-cause mortality rate, ICU admission rate and length of hospitalisation (LOS) across these *K. pneumoniae* types, identified genome-wide determinants linked with clinical severity and used machine learning approaches to predict clinical severity outcomes from genomic biomarkers together with clinical metadata.

**Results:** Infections caused by convergent strains exhibited the greatest clinical severity, showing nearly double the in-hospital mortality (reaching 42% at 90 days), a 2.4-fold higher likelihood of ICU admission, and an average 150% increase in LOS compared to infections caused by susceptible and non-hypervirulent strains. Our findings indicate an additive effect of hypervirulence and multidrug resistance on disease severity. Carbapenem resistance determinants showed the strongest association with adverse outcomes, even after adjusting for the presentce of other resistance and virulence genes and clinical confounder features. The GWAS analysis revealed associations of the clinical outcomes with accessory genes involved in carbohydrate metabolism and the Type VI secretion system (T6SS) machinery, metabolic-adaptation and stress-tolerance/persistence loci. Additional significant associations were identified with SNPs in ABC-transporters, cell-envelope systems, sugar transporter families and RND-family efflux systems. Machine-learning models yielded average Area Under the Curve (AUC) values of 0.78 and 0.79 for mortality and ICU admission, respectively, and exhibited strong monotonic association between observed and predicted outcomes for LOS, with an average correlation of 0.59 on unseen test data when trained using combined genomic and clinical predictors.

**Conclusion:** This study identifies key genomic determinants that drive severe *K. pneumoniae* infections, with carbapenem-resistance markers emerging as the leading contributors to poor clinical outcomes. The strong predictive performance of genomic biomarkers, particularly for mortality, ICU admission, and LOS, highlights their value in enhancing diagnostic precision, improving clinical risk stratification, and informing targeted infection-prevention strategies.

## Introduction

*Klebsiella pneumoniae* (KP) is a major Gram-negative bacterial species that is commonly found in the normal human gut microbiota. Although often commensal, KP is an opportunistic pathogen capable of causing severe infections including pneumonia, bloodstream infections, intra-abdominal infections and septic shock [1–4]. Among these, resistant KP, and carbapenemase-producing KP (CP-KP), driven in large part due to extensive antimicrobial use, are now among the most frequent pathogens recovered from hospital-acquired infections worldwide [5, 6]. In economic terms, CP-KP infections generated an estimated $4.46 billion in socioeconomic costs in 2019 alone, with losses predominantly driven by productivity impacts rather than direct healthcare spending [7]. Transmission of these strains, associated with significantly worse clinical outcomes compared to susceptible strains, is primarily nosocomial [8–10]. In parallel, hypervirulent KP (hvKP) has emerged as a distinct pathotype characterised by enhanced virulence and the ability to cause life-threatening community-acquired infections such as liver abscesses and sepsis [11]. More recently, the convergence of multidrug-resistance and hypervirulence has emerged as an alarming threat. Specifically, strains harboring both third-generation cephalosporin (3GC) resistance, producing ESBLs, or carbapenem resistance, and virulence determinants, often acquired via plasmid transfer or hybrid plasmid formation, have been documented across multiple geographies. These “superpathogen” strains combine high-level antimicrobial resistance with hypervirulence, significantly increasing their capacity to cause severe, hard-to-treat infections [12, 13]. Clinically, these convergent combines the detrimental features of both pathotypes, resulting in severe infection, frequent outbreaks and exceptionally high mortality rates.

Since genomic definitions of 3GC resistant (ESBL(+)), CP(+) and hv phenotypes have become more accurate and clinically relevant, an increasing number of studies have sought to characterize their clinical manifestations, risk factors, and mortality. Across multiple large-scale analyses, infections caused by ESBL(+) and CP(+) strains clearly exhibit substantially worse outcomes than those caused by susceptible strains [14–16]. A meta-analysis found pooled mortality rates of approximately 42% for CP-KP versus 21% for susceptible strains [17]. Additional studies show that CPKP infections are associated with increased ICU admission and higher costs of care [18, 19]. Genomically defined hvKP, defined based on the presence of hallmark virulence genes, is associated with increased early mortality in matched patient cohorts [20]. The combined effect of ESBL(+) or CP(+) and hypervirulence on infection severity is, however, inconsistent: although convergent outbreaks (often ST11 KL64) display extremely high unadjusted lethality particularly in ICU settings [21, 22], other studies within carbapenem-resistant strata report that hv-CPKP does not consistently result in worse outcomes compared to classical CP-KP and ESBL-KP [4]. This likely reflects variation in infection sources, patient populations, and study designs.

Despite growing evidence of the clinical significance convergent strains (i.e. hv-CRKP / hv-ESBL-KP), prior studies have almost exclusively assessed the effects of resistance and virulence in isolation, without quantifying their combined impact. These studies have primarily focused on mortality endpoints, while critical dimensions such as ICU admission and infection-attributable length of stay (LOS) remain largely unexamined or inconsistently reported. These outcomes require standardized, adjusted measures and prospective capture. Moreover, previous studies have mainly examined panels of known resistance and virulence determinants in relation to clinical severity outcomes; consequently, potential genome-wide causative and predictive biomarkers have not been identified. Finally, although several reports have linked virulence and resistance genotypes to adverse outcomes, the predictive and prognostic value of these genetic determinants for mortality and disease severity has not been systematically evaluated in large cohorts. Addressing these limitations will be crucial for refining risk models and improving the clinical translation of genomic data.

To address these gaps, we analysed a large-scale, whole-genome collection of the *K. pneumoniae* complex, systematically obtained through a multicentre, nationwide framework over seven years across the Kingdom of Saudi Arabia (KSA). We integrated whole-genome sequencing data with detailed patient history and clinical metadata to identify genomic determinants and risk factors associated with clinical severity. We aimed to identify genetic determinants and risk factors associated with infection and to evaluate the clinical outcomes of infections caused by strains carrying known convergent KP markers. Through bacterial genome-wide association studies (bGWAS) and multivariate analyses, we quantified the association of genomic variants with in-hospital mortality, ICU admission, and LOS and identified novel biomarkers for these outcomes. We then integrated the biomarkers in a machine-learning framework and demonstrated that bacterial genomic biomarkers harboured significant predictive power for adverse clinical outcomes. Together, these findings provide a comprehensive integration of convergent *K. pneumoniae* genotypes with patient-level clinical data and highlight the clinical utility of genomic biomarkers for severity prediction.

## Methods

### Sampling, short-read sequencing and epidemiological cluster analysis

We provided details on ethical approvals, sampling approach, and antimicrobial susceptibility testing in the Supplementary Methods. In brief, we subjected a total of 1,178 non-duplicate clinical isolates of the *K. pneumoniae* complex, collected from five nationwide hospital centers between 2018 and 2025, for short-read whole-genome sequencing. The quality of the sequencing reads was assessed using the FastQC package in R (v0.1.3). We assembled the genomes *de novo* with Unicycler (v0.5.0) using default settings, and discarded contigs under 200 bp from subsequent analysis [23]. We annotated assemblies using Bakta (1.8.2) [24] and resulting assemblies were fed into Panaroo for pangenome reconstruction [25]. Resistome and virolome characterization and MLST typing were carried out with Kleborate (v2.3.2) to assign species, sequence type, K-locus, and to profile resistome and virulome [26]. We used the gene-cased scoring scheme of Kleborate to determine the virulence and resistance levels and to identify hypervirulent and carbapenem and cephalosporins resistant and covergent isolates. The computed virulence (0–5) and resistance (0–3) scores were derived using the following scheme. For virulence: yersiniabactin (*ybt*) < colibactin (*clb*) < aerobactin (*iuc*). Scoring: 0 = none of *ybt/clb/iuc*; 1 = *ybt* only; 2 = *ybt* + *clb* or *clb* only; 3 = *iuc* only; 4 *= iuc + ybt* without clb; 5 = *ybt + clb + iuc*. For resistance: ESBL < carbapenemase < carbapenemase + colistin. Scoring: 0 = no ESBL or carbapenemase; 1 = ESBL only; 2 = carbapenemase without colistin resistance determinants; 3 = carbapenemase plus colistin resistance determinants. Based on these scores, we defined four pathotypes. Strains with a resistance score ≥ 1 were grouped as ESBL(+)/CP(+), i.e. ESBL-producing (ESBL(+)) or carbapenemase-producing (CP(+)). Genomes with a virulence score ≥ 3 as hypervirulent (hv) [26]. Convergent strains were defined as genomes with resistance score ≥ 1 and virulence score ≥ 3 [26]. The resistome and virolome profile of Kleborate was used in the downstream bacterial GWAS and machine learning predictive modelling. The resistome and virolome profiles were also used in the downstream multivariate analyses and machine learning predicitive modelling as input features.

To identify single-nucleotide polymorphism (SNP) in the core genome, we aligned short-read data against the genome of the reference strain *K. pneumoniae* Ecl8 [27] (ST375-K2; accession PRJEB401) using the Snippy pipeline with default parameters. Pair-wise SNP distances were then calculated, and a neighbor-joining tree was built in R using the ape package (v5.7.1) [28]. We used the annotated output of Snippy of SNPs for downstream analysis and filtering missense and stop-gain mutations. We also leveraged the sequencing data to identify epidemiological clusters across all lineages in the collection; comprehensive methodological details are provided in the Supplementary Methods

### Multivariate regression analysis

We examined the association between the presence of genetic determinants and strain pathotype (hv status and ESBL/CP status) with three clinical severity outcomes, i.e. in-hospital all-cause mortality (discharged alive vs. deceased), ICU admission (ICU admitted vs. not-admitted) and length of hospitalization (LOS), in the visit, where the infection was diagnosed. We developed three analytical frameworks to evaluate the clinical effect of genomic determinants under different conditions: first, to assess the effect of each determinant in isolation from other determinants (Model (M)1); second, to examine the joint effect of resistance and virulenc determinants in the same model (M2); and third, to evaluate the effect of having both resistanc and virulence determinants, as well as the potential interaction between resistance and virulence levels (M3). In all three frameworks, patient-level clinical data were included to adjust for confounding.

In all modelling frameworks, for ICU admission and in-hospital mortality, we modeled the binary outcomes (ICU admitted vs. not admitted; deceased vs. alive) using multivariable logistic regression to estimate the association between exposure to each genotype in isolation from other genes/mutations. For LOS, which is a continuous variable with a right-skewed distribution, we fitted a multivariable linear regression model after applying a log(LOS + 1) transformation to reduce skewness and stabilize variance.

The first modelling framework (M1) for ICU admission and mortality is the following logistic regression model:

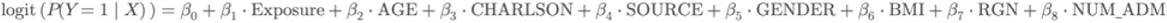

where *Y* represents the clinical outcome and can be ICU admission, or in-hospital death. Exposure represents the genetic profile of the infecting isolate, defined as the presence or absence of virulence and/or resistance genes or mutations identified by *Kleborate*, the pan-genome, the SNP-calling pipelines or their pathotypes (i.e. hypervirulence or resistance), a defined by Kleborate. The other terms represent clinical patient level data as clinical covariates. This includes Age, Charlson comorbidity index, and BMI represent the patient’s clinical characteristics during hospitalization, while source refers to the anatomical site of infection (blood, urine, respiratory, or other specimen types). RGN refers to the hospital region, and NUM_ADM denotes the number of the patient’s admissions prior to the infection. For LOS, we fitted a linear regression model using the same set of confounders.

Because many genetic features are highly correlated owing to the co-occurrence of multidrug-resistant and hypervirulent determinants on the same genetic background, we also developed models (M2) that included all genes simultaneously to assess the effects of individual genes in the context of co-occurring determinants, using following formula:

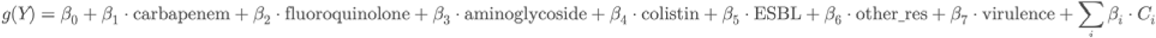

In this formula, *g*(.) denotes the link function for the outcome being modeled: an identity link when *Y* represents length of stay (linear regression), and a logit link when *Y* corresponds to ICU admission or in-hospital mortality (logistic regression). The coefficients *β*_1_ through *β*_7_ quantify the association between each antimicrobial-resistance class or virulence determinant, a determined by Kleborate, and the clinical outcome. The term Σ*_i_ β_i_*, *C_i_* captures the abovementioned covariates, , which are summarized collectively. In this model, other_res denotes resistance determinants against antimicrobial classes other than aminoglycosides, colistin, carbapenems, and cephalosporins (ESBL-producing enzymes), capturing resistance to all remaining drug categories not included above.

We quantified multicollinearity using the Variance Inflation Factor (VIF) computed via the vif() function in the car R package. We applied a conservative cutoff of VIF < 5 for non-problematic collinearity. This ensured that coefficient estimates were not inflated by redundant predictors and that the independent effects of each resistance class and virulence score could be interpreted reliably [29].

Finally we developed model 3 (M3) to assess the effect of interactions between resistance and virulence determinants on clinical severity features:

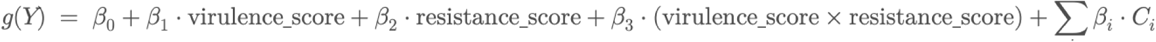

In this formula *g*(.) and Σ*_i_ β_i_*, *C_i_* are defined as above. Th term *β*_1,_ *β*_2_ are the main effects of virulence and resistance levels as quantified by resistance and virulence scores, *β*_3_ is the interaction effect between virulence and resistance levels.

### Survival analysis

We performed survival analysis (with the survival and survminer R packages) to examine the association between exposure to pathotype classes (hv and ESBL/CP gene combinations) and in-hospital mortality. First, we employed Kaplan–Meier model in which survival time was defined as length of hospitalization stay, and in-hospital mortality as a binary indicator (1 = death, 0 = censored). We then stratified exposures to isolates into four groups: “Others”, “Only hv”, “Only ESBL(+)/CP (+)”, and “Convergent” based on virulence and resistance scores. We fitted stratified survival curves Surv(Survival Time, Death Event) ∼ Exposure Type, including p-value and 95% confidence intervals to compare survival among genotype groups. In additional analyses, we fitted a multivariable Cox proportional-hazards model to estimate the association between time to event (length of stay, LOS) and in-hospital mortality while adjusting for clinical confounders. We defined the time origin as hospital admission (time zero). Follow-up ended at in-hospital mortality (event) or discharge alive (censoring). LOS from admission to discharge or death was used as the time-to-event variable; if no death occurred by discharge, the patient wa censored at the discharge date. We used the following model:

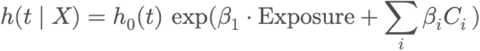

where *h*(*t* | *X*) is the hazard at time *t* given covariates , and *h*_0_(*t*) is the baseline hazard. The term Σ*_i_*, *β_i_C_i_* captures the abovementioned clinical covariates.

### Long read sequencing and plasmidome analysis

To resolve the full plasmidome content of the collection, we selected 179 isolates from each sequence type (ST) that were represented by more than three genomes. Isolates exhibited a unique pattern of ESBL or carbapenemase plasmid-borne virulence genes, as reported by Kleborate. Plasmid DNA was isolated from 4 ml overnight cultures grown in LB medium using the QIAprep® Miniprep Kit following the manufacturer’s protocol. We assessed plasmid concentration via Qubit HS dsDNA assays and determined purity Determined purity using a nanodrop. For sequencing, libraries were prepared and multiplexed using 96-plex Rapid Barcoding Kits, then loaded into Oxford Nanopore MinION flow cells and run for 48h according to the manufacturer’s instructions. Hybrid assembly was subsequently performed using Unicycler with the conservative option to generate plasmid assemblies. We identified putative plasmid fragments based on the presence of a known plasmid replicons reported by abricate (v1.0.1) (www.github.com/tseemann/abricate) and filtered these fragments with Bandage (v0.9.0) [30]. We employed Kleborate to determine resistance and virulence determinants/scores for each plasmid. We linked plasmid carriage status with clinical severity outcomes by computing the Relative Risk (RR) for the binary outcomes of in-hospital mortality and ICU admission and the Rate Ratio for the continuous log-transformed LOS, using modelling settings similar to Model 1 with adjustment for clinical confounders. In this context, RR quantifies the probability of a clinical event in one plasmid exposure group relative to another, allowing comparison of risk between plasmid-defined exposure subgroups in our cohort.

### Bacterial genome wide association studies (bGWAS) analysis

To systematically identify genetic elements associated with three clinical features—namely hospital mortality, ICU admission and LOS—we conducted bacterial genome-wide association studies (bGWAS) using binarized outcome features. ICU admission and in-hospital mortality were already treated as binary variables. To LOS labels, hospitalization days were log - transformed and then dichotomized at the cohort median to represent short versus long stays. We then performed GWAS using these dependent features and genomic predictors, which included presence/absence of accessory genes, defined by Panaroo, and SNPs identified using Snippy. We fed the data into the Scoary (v.1.6.16) pipeline [31] with 50 iterations to quantify association strength and significance between binary-genotype features and phenotypic traits.

Scoary first runs a Fisher’s exact test for each gene-trait pair and applies a Benjamini–Hochberg correction for multiple testing. Then, for features passing the initial threshold, it uses a phylogeny-based algorithm to identify the maximum number of non-intersecting isolate pairs that differ in both gene presence and trait status, and reports both the best (lowest) and worst (highest) possible p-values from these comparisons, thereby reducing lineage-confounding effects. In our workflow, we filtered genes that met the criteria of *p* < 0.05 in the naïve test and passed thresholds on BH-adjusted *p*-values (< 0.05). We then further restricted those genes by pairwise comparison criteria: for strict associations, both best and worst pairwise *p*-values had to be < 0.05, while for broader associations only the best pairwise *p*-value needed to be < 0.05.

After identifying significant accessory-gene families, we performed BLAST searches to verify their presence and exclude artificial associations due to gene fragmentation. Using the consensus gene sequences reported by Panaroo, we confirmed gene presence in the assemblies and removed any associations not resulting from genuine gene presence. We then conducted regression analyses, as detailed in Model 1, to retain only those genes/SNPs whose presence remained positivly associated with the clinical outcomes after adjusting for clinical confounders (Z-test p-value < 0.05). After identifying gene families linked with in-hospital mortality, we excluded hypothetical gene families and applied *k*-means clustering, using the elbow method to select the cluster number, to group the hit genes based on their co-occurrence. Full GWAS results are provided in the GitHub directory of the project (www.github.com/hjy1805/Kp_multicentre_hospitals).

### Machine learning framework

We developed a machine-learning based toolkit to predict clinical outcomes of bacterial infections using genomic biomarkers. The response features included binarized in-hospital mortality and ICU admission (for which classifier models were employed) and a continuous LOS feature for which regressor models were used. The predictive framework incorporated both baseline regression and ensemble models, trained, validated, and evaluated using integrated genomic and clinical datasets. The clinical and demographic variables included age, body mass index (BMI), Charlson comorbidity index, region, and body source of infection.

Genomic biomarkers included antimicrobial resistance and virulence genes identified using Kleborate, the pangenome defined by Panaroo, SNPs called via Snippy, and unitigs (i.e., unique sequence fragments formed by merging kmers into maximal unambiguous stretches) generated by the unitig-caller pipeline (www.github.com/bacpop/unitig-caller) [32]. We employed Scoary for feature-engineering to reduce the predictor set. For pangenome, SNP, and unitig data, we filtered variants with BH-adjusted p-values < 0.05. After filtering, the final feature sets included 20,294 SNPs and 2,715 accessory-genome features and 14,128 unitigs for the mortality model; 5,468 SNPs, 5,752 unitigs, and 2,147 accessory-genome features for the ICU-admission model; and 11,619 SNPs, 12,764 unitigs, and 2,365 accessory-genome features for the LOS model.

We also conducted One Hot encoding on the cathegorical clinical predictor features to convert them into numerical features. We used the filtered features as predictors and trained models on progressively richer datasets. The first dataset contained only clinical and demographic variables; subsequent datasets incrementally incorporated antimicrobial resistance and virulence determinants, pangenome content, SNPs, and unitigs, resulting in five levels of training data. To examine the prediction information in genomic data, we used all genomic data predictors in isolation from clinical data as predictor as well.

Training data were partitioned into 80% training/validation and 20% unseen test sets in five non-overlapping folds. Within the training set, we applied three-fold cross-validation and hyperparameter optimization through grid search. Two models were evaluated for predicting ICU admission and in-hospital mortality: a baseline elastic net classifier and an ensemble XGBoost model [33]. For the prediction of LOS, a continuous outcome, we employed models that not only provide point estimates but also generate prediction intervals, thereby quantifying uncertainty in the predictions and accounting for the nature of the dependent variable. The models included a baseline linear quantile regression model and the NGBoost [34] framework for probabilistic and interval prediction. NGBoost estimates full conditional distributions rather than single-point predictions by modeling parameters such as the mean and variance within a multi-parameter boosting framework.

For the gradient-boosting XGBoost models, we tuned hyper-parameters including tree depth (2 or 3), sub-sampling ratio (0.8 or 1), number of estimators (2000), and learning rate (0.03). For the NGboost model, hyper-parameters included tree depth (2 or 3), number of estimators (100, 200), and learning rate (0.03). For the elastic net model, we tested two levels of regularization strength (C = 0.1 and 1.0), covering pure ridge, balanced elastic-net, and pure lasso penalties, and three penalty types (l1_ratio = 0.0, 0.5, 1.0, ranging from strict to more lenient) to see which combination gave the most reliable predictions. For the quantile regressor model we tested a range of “shrinkage” values (α= 10^−4^, 10^−3^, 10^−2^, 0.1, 1.0) to prevent overfitting. AUC and MAE was used to select the best-performing model for classifiers and regressors, respectively.

Model performance was evaluated using the area under the receiver operating characteristic curve (AUROC), precision, recall, and F1-score for classification models, and the coefficient of determination (R²), the correlation between predicted and observed values on the test set, and the coverage of 90% prediction intervals to assess uncertainty estimation for quantile prediction models. Feature importance and feature interactions were quantified using SHAP (SHapley Additive exPlanations) values [35] computed within each cross-validation fold and aggregated across folds to identify features consistently predictive of outcomes. SHAP values, derived from cooperative game theory, provide a unified measure of feature contribution by estimating how each variable (i.e., gene, virulence factor, or clinical attribute) influences individual model predictions. Feature interactions with SHAP was assessed by computing interaction values, which quantify how much the joint effect of two features on model predictions differs from the sum of their individual contributions. Assessing these interactions informs on complex, non-additive relationships between predcitors. To reliably identify interactions, we repeated the predictions ten times on random splits of the training and test datasets and retained only those interactions that appeared consistently across runs. This approach helps ensure that identified interactions reflect stable model behavior rather than random variation due to a specific data split. The code, details of the model implementation and intermediate files are available in the project’s GitHub repository (https://github.com/hjy1805/Kp_multicentre_hospitals). We also deployed the trained model as a standalone Streamlit application, which users can run locally (www.github.com/hjy1805/Kp_prediction).

## Results

### Prevalnce of ESBL(+)/CP(+) and hypervirulent clones across the hospitals

We analyzed a genomic collection of 1,306 *K. pneumoniae* complex isolates retrieved from 2018 to 2025 in a nationwide hospital network. The collection was highly diverse, encompassing 374 distinct sequence types (STs), of which 18 major STs (each represented by more than ten isolates) together account for 45 % of the total population (Figure S1A). The frequency of clones exhibiting both hypervirulence (hv) and ESBL(+)/CP(+) status was approximately 35% of the total isolates and remained stable throughout the duration of the study (Figure S1B). Integrating resistance and virulence scores with clone (ST) frequencies revealed a strong positive correlation (p value < 0.0001 from Pearson correlation test) with both scores, suggesting that higher resistance and/or virulence levels drive increased clone prevalence (Figure 1 and S1A). Predominant clones also exhibited higher resistance levels to carbapenems and cephalosporins (Figure S2A). Among the most prevalent clones were those known to be emerging or globally circulating. Most notably, ST2096-KL64, comprising approximately 15% of our collection (Figure 1), was previously documented as a regionally expanding convergent clone in Saudi Arabia and broader South/West Asia and showed the highest rate of carbapenem resistance [36, 37] (Figure S1, S2A). The collection also included well-characterised ESBL(+)/CP(+) clones of ST101, ST11 (part of CG258), ST14, ST307 (CG307), and ST37, all exhibiting carbapenem resistance (Figure 1, S1A and S2A). Within the major ESBL(+)/CP(+) clones of ST11-KL47, ST147-(KL64/KL10) and ST101-KL17, genomes sporadically harboured also hypervirulence genes, classifying them as convergent strains (Figure S2). Carbapenem-resistance in the collection was predominantly driven by *bla*_OXA-48_-like enzymes, i.e. *bla*_OXA-48_, *bla*_OXA-232_ and *bla*_OXA-181_ in ST2096, ST147, ST231 and ST101, while *bla*_NDM-1_ and *bla*_NDM-5_ occurred across multiple STs including ST147 and ST11. *bla*_KPC-2_ was largely restricted to the ST11 lineage. Porin mutations (i.e., OmpK36 loop-3 insertions (GD/TD) and OmpK35 truncation/deletion) and colistin resistance mutations, mainly in *mgrB* genes, and less frequently in *pmrB*, co-occurred in the ESBL(+)/CP(+) clones of ST101, ST11, ST14, ST147 and ST2096, consistent with synergy between carbapenemases and porin loss (Figure S2). Variants of the *bla*_CTX-M_ gene represented the most common ESBL genes in the major clones and also in less-frequent clones (i.e., ST866, ST380, ST268, ST592) (Figure S2A). The collection also comprised hypervirulent clones, including the canonical ST23-KL2 clone and the emerging ST268-KL1 and ST86-KL2 clones, each harbouring hallmark virulence genes for salmochelin (*iro*) and aerobactin (*iuc*) and all displaying low resistance levels (Figure S2B). In addition, rare novel hv clones of ST2846, ST1970 and ST382 were identified. While the frequency of dominant clones remained constant over the eight-year study period (Figure S1B), the largest clones of ST2096 and ST147, exhibited a significant increase in resistance scores over time (pvalue < 0.05, trend test). This was driven by higher frequencies of *bla*_OXA-48_-like and *bla*_NDM-1_ carbapenemases in ST2096 and ST147 clones, respectively. The ST147 clone also exhibited a marked increase in virulence, driven by the acquisition of the aerobactin (iuc) locus, indicating ongoing evolutionary diversification of this lineage. Beyond intra-clone evolution and the rise of hv lineages, our data reveal a diverse yet stable *K. pneumoniae* population circulating nationwide over seven years.

**Figure 1.**
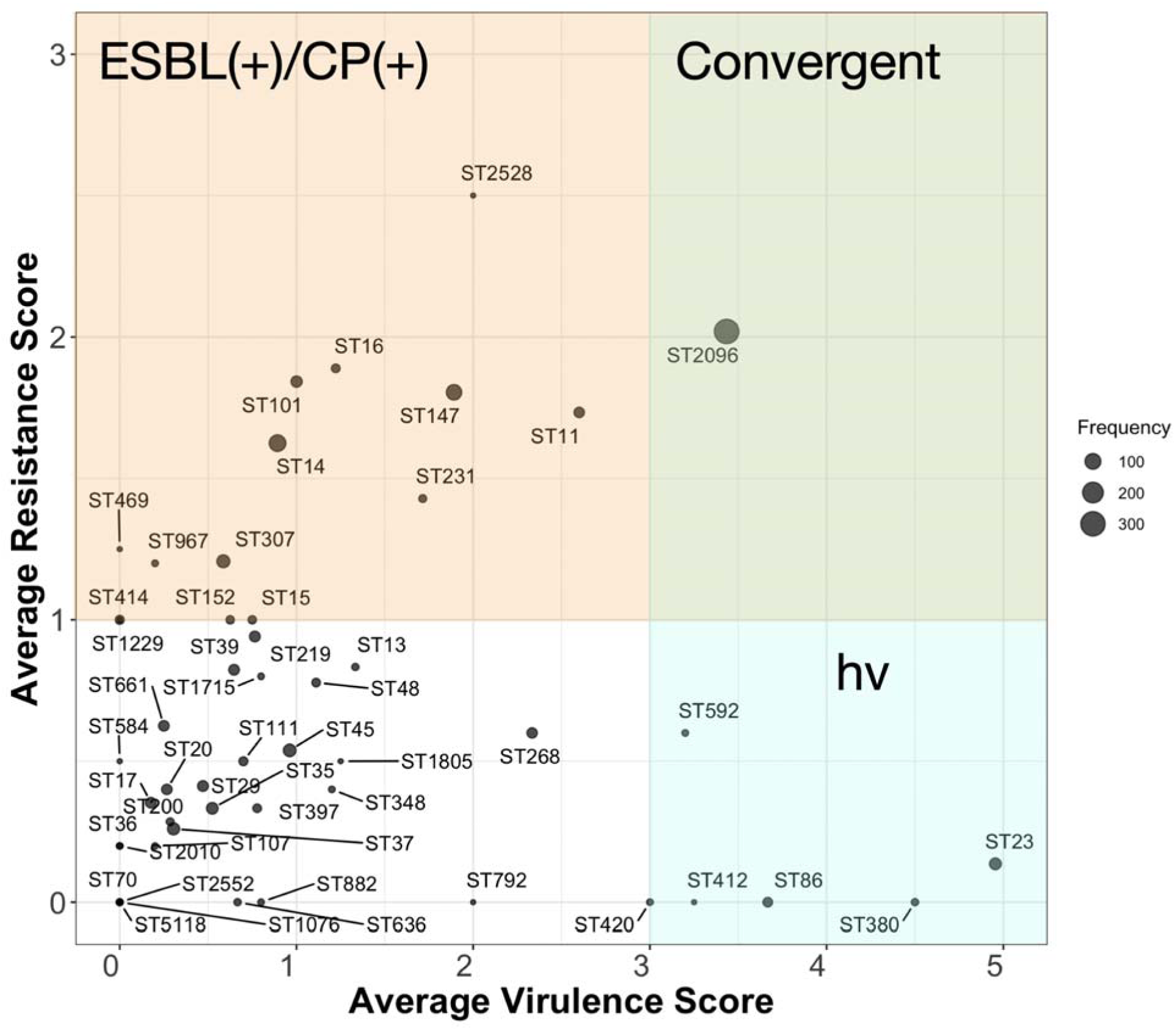
The frequency of sequence types (STs) based on their resistance and virulence category. The size of each bubble corresponds to the frequency of the isolates with the particular sequence type. The x and y coordinates denote the average resistance and virulence scores for the isolates within the ST. Hypervirulent (hv) and ESBL(+)/CP(+) states were defined by Kleborate scoring schemes. Resistance scores were assigned based on the presence of major antimicrobial resistance determinants as follows: 0 = no ESBL or CP genes detected, 1 = ESBL(+)/CP(-), 2 = CP(+), and 3 = CP(+) + colistin resistance mutations (*mgrB/pmrB*) or genes. Virulence score were calculated according to the presence of key virulence loci: 0 = none, 1 = *ybt* (yersiniabactin), 2 = *ybt* + *clb* (colibactin), 3 = *iuc* (aerobactin), 4 = *ybt* + *iuc* and 5 = *ybt + clb + iuc*.

### Over-representation of convergent clones within the epidemiological clusters in the hospital network

Clustering of genomes from hospitals using a transmission cutoff revealed 145 distinct transmission clusters, which were strongly associated with both pathotypes and clincial characteristics of the isolates (Figure 2A). The clusters included 268 isolates in total representing 23% of total isolates with full clinical metadata. While non-hv non-resistant strains were mainly scattered and rarely clustered, ESBL(+)/CP(+) and convergent isolates frequently formed large connected groups. Clusters were distributed across hospital regions, with some extending across multiple sites, and persisted over several years, highlighting both regional spread and long-term circulation of dominant clones (Figure 2A). The largest clusters corresponded to the convergent ST2096 and ESBL(+)/CP(+) ST14, as reported as cirulating clones in the country for country-wide study in 2022/2023 [37]. The networks suggest significant nosocomial spread. Isolates from hospital-acquired infections were about twice as likely to belong to transmission clusters (OR:2.0, 95% CI 1.3–3.0), whereas community-acquired isolates were markedly less likely to cluster (OR:0.5, 95% CI 0.3–0.8) (Figure 2B). Moreover, overlapping hospitalization stay periods were a strong predictor of clustering, with particularly high odds observed when patients were admitted to or discharged from the same ward (OR: 6.0–8.1), and the effect was most pronounced for direct overlap within the same ward and hospitalisation stay (OR:20, 95% CI: 7.0–32.2) (Figure 2B). Convergent isolates had the highest odds of clustering (OR:8.0, 95% CI: 5.5–11.2), followed by ESBL(+)/CP(+) only isolates (OR:1.95, 95% CI: 1.5–2.3). By contrast, hypervirulent isolates had significantly lower odds of belonging to a transmission cluster (OR < 1), indicating they are more likely to occur as sporadic cases rather than part of sustained hospital transmission clusters (Figure 2B). This pattern remained significant after accounting for hospitlisation length of the patients, indicating that the excess clustering observed among ESBL(+)/CP(+) and convergent isolates is driven by their enhanced capacity for nosocomial transmission and persistence within hospital environments, rather than prolonged patient exposure. This widespread intra- and inter-hospital transmission of resistant clones is consistent with the documented dissemination of globally prevalent high-risk lineages, such as ST258 and ST11, across healthcare networks [38, 39].

**Figure 2.**
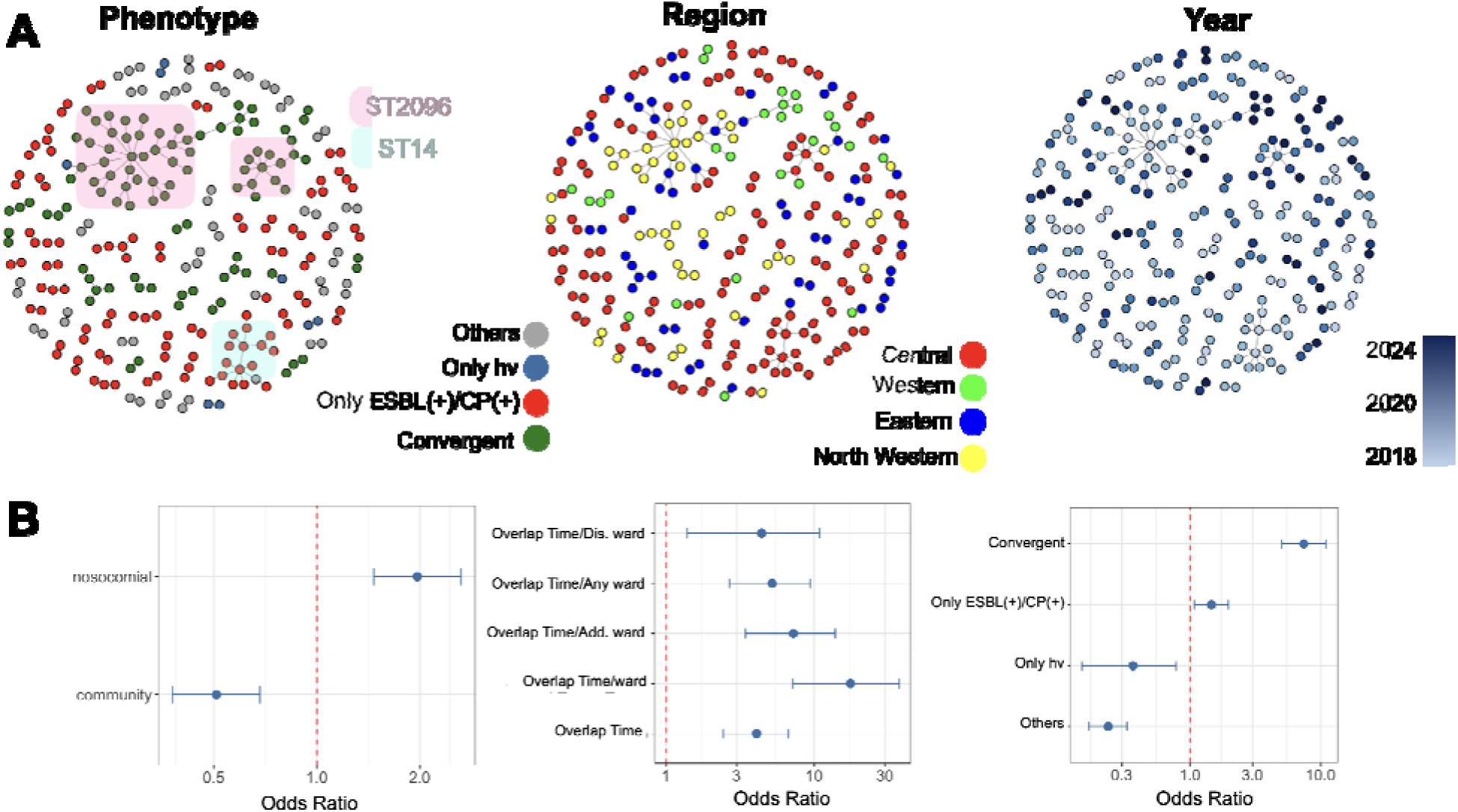
Epidemiological cluster analysis. **A)** Identified networks corresponding to the SNP cutoff of 20 for the isolates infecting distinct patients. Each node represents one strain from one patient. Nodes represent potential epidemiological links (see Methods). The phenotype subpanel shows the pathogencity and drug resistance level of strains based on the scores inferred from Kleborate pipeline. Region and year are based on the region of the hospital of origin and the year of isolation. B) The association of infection and strain types with the cluster status of isolates. Nosocomial infections are strains recovered within 48 hours of the patient’s admission to the hospital, while community infections are strains recovered beyond 48 hours of the patient’s hospitalization. Overlapping time indicates overlapping hospitalisation length for the stay during which the infection was diagnosed. “Overlapping ward” indicates that patients were in the same admission (add.) or discharge (dist.) ward, or both. Error bars denote 95 % confidence intervals.

### Significant impact of convergent status on in-hospital mortality

In addition to their higher odds of clustering, both hv and ESBL/CP status were strongly linked with elevated in-hospital mortality (p < 0.005) (Figure 3A) and they likewise corresponded to higher 30-, 60- and 90-day mortality rates (Figure 4A). While both hv and ESBL/CP status were associated with increased 30-day mortality (21% higher than baseline), the effect of ESBL(+)/CP(+) alone (8% above baseline) clearly exceeded that of hv (5% above baseline), underscoring that ESBL(+)/CP(+) status exerts a stronger influence on fatal outcomes than hypervirulence (Figure 4A). The results from the survival analysis also demonstrated significant differences in patient outcomes across the exposure groups to four *K. pneumoniae* pathotypes, when stratified by length of stay (LOS) (Figure 4B, p < 0.0001) a pattern that became evident as early as 10 days of hospitalization (Figure 4B). Multivariable Cox regression analysis, which included the effects of potential confounders, further confirmed these associations (Figure 4C). Compared with non-hv ESBL(-)/CP(-) infections, ESBL(+)/CP(+) infections were significantly associated with increased mortality risk (Hazard Ratio (HR) 1.45, 95 % CI 1.16–1.81, p < 0.001), while convergent infections carried the highest hazard, nearly doubling the risk of in-hospital mortality (HR 1.55, 95 % CI 1.17–2.04, p < 0.001) and showing half hazard of bloodstream infections (Figures 4B, 4C). Although survival in the hv only group was not significantly different from that of the non-hv ESBL(–)/CP(–) group, the presence of hypervirulence determinants combined with ESBL/CP determinants appeared to contribute to in-hospital mortality, indicating that the convergence of hypervirulence and multidrug resistance is the most critical driver of poor outcomes in this cohort.

**Figure 3.**
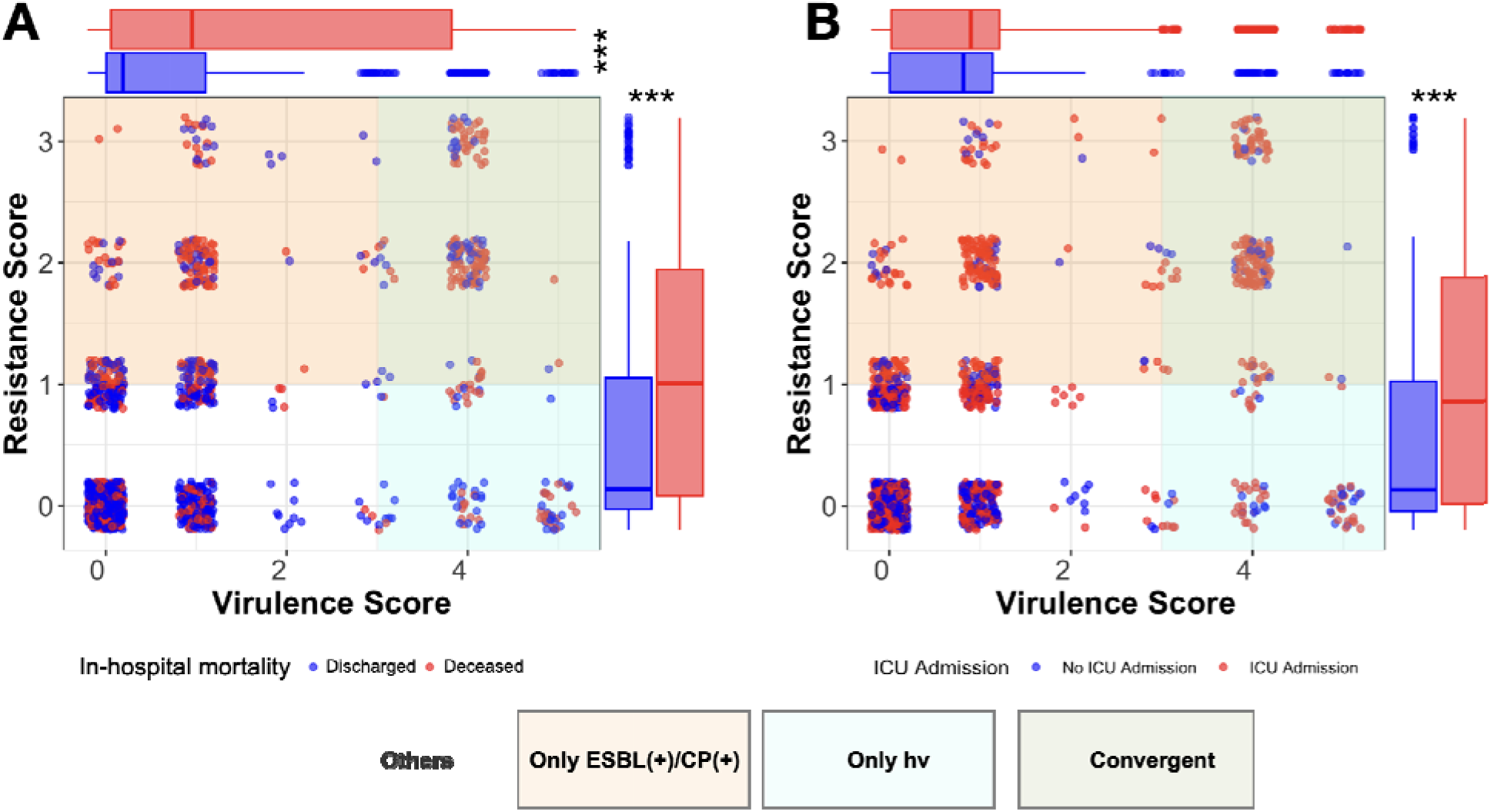
The association of virulence and resistance score with ICU admission and in-hospital mortality rates. Each dot represents one isolate, with the color representing the status of the patient from whom the isolate was recovered. The colors red and blue correspond to deceased/ICU-admitted and discharged/no ICU admission in the mortality/ICU admission plots. The boxplots are an aggregated distribution of resistance and virulence scores. The asterisks denote the significance of the difference in the mean for the aggregated resistance and virulence score plots from the Wilcoxon rank-sum test, with *** indicating a significance level of < 0.001.

**Figure 4.**
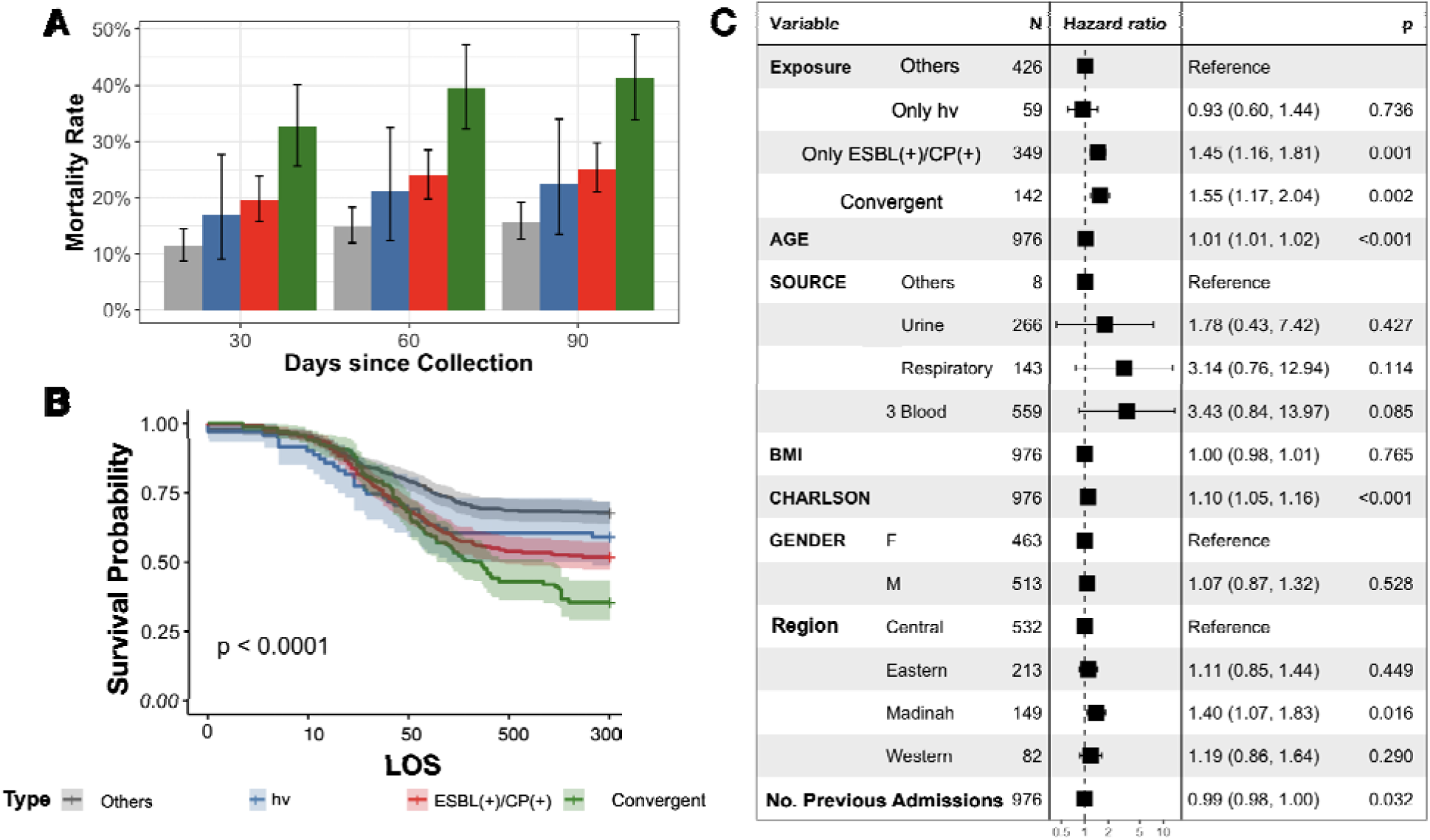
Mortality and survival analysis for the patients based on the risk group. **A)** mortality rates based on the date of death from the day of sample collection. **B)** Kaplan-Meier plots for patients under study based on the resistance and virulence category of the isolate. The ribbon area denotes a 95% confidence interval. The p-value is from the Mantel–Cox test between at least two of the four groups. C) Multivariable Cox proportional-hazards regression anlysis, which models how each covariate affects the mortality hazard after adjusting for all other variables in the table. The p-values in C) is from Wald tests within a Cox regression, quantifying the statistical significance of each variable’s adjusted hazard ratio.

### Increased ICU admission rates and extended hospitalisation in infections caused by convergent srains

Our results indicate a mostly similar effect of infections by hv and ESBL(+)/CP(+) pathotypes and additional clinical severity features of ICU admission and hospital length of stay (LOS). Although both of these severity outcomes correlate with in-hospital mortality, the correlations were weak (r = 0.29 95% CI: 0.23-0.34 for ICU admission and r = 0.21 95% CI: 0.16-0.27 for LOS). Consequently, we examined the associations between exposure to specific pathotypes and each of these severity outcomes separately to dechiper their distinct impacts. Consistent with the patterns observed for in-hospital mortality, ICU admission was strongly associated with antimicrobial resistance status, with significantly increased odds among MDR non-hypervirulent isolates (OR = 1.77, 95% CI 1.33–2.36) and the highest risk observed for hv-MDR isolates (OR = 2.45, 95% CI 1.64–3.66). In contrast, hypervirulence in the absence of resistance was not significantly associated with ICU admission (OR = 1.23, 95% CI 0.72–2.09), indicating that resistance rather than virulence is the primary driver of ICU-level disease severity (Figure 3B). A similar pattern was observed for the association with LOS, both with (Figure 5A) and without adjustment for confounding factors (Figure 5B). While infections caused by hv only strains were not linked to longer hospital stays, ESBL(+)/CP(+) status alone was associated with significantly longer mean stays, and when combined with hv status the effect was even more pronounced, showing about 80 days (CI: 48-120) longer for ESBL(+)/CP(+) strains, and around 150 days (CI: 85-220) longer for convergent strains compared with non-hv ESBL(–)/CP(–) exposure (Figure 5A, 5B). Altogether, these results indicate that hvKp strains do not independently account for prolonged hospitalization, length-of-stay–stratified in-hospital mortality, or higher ICU admission rates in hospital settings when dissociated from ESBL/CP determinants.

**Figure 5.**
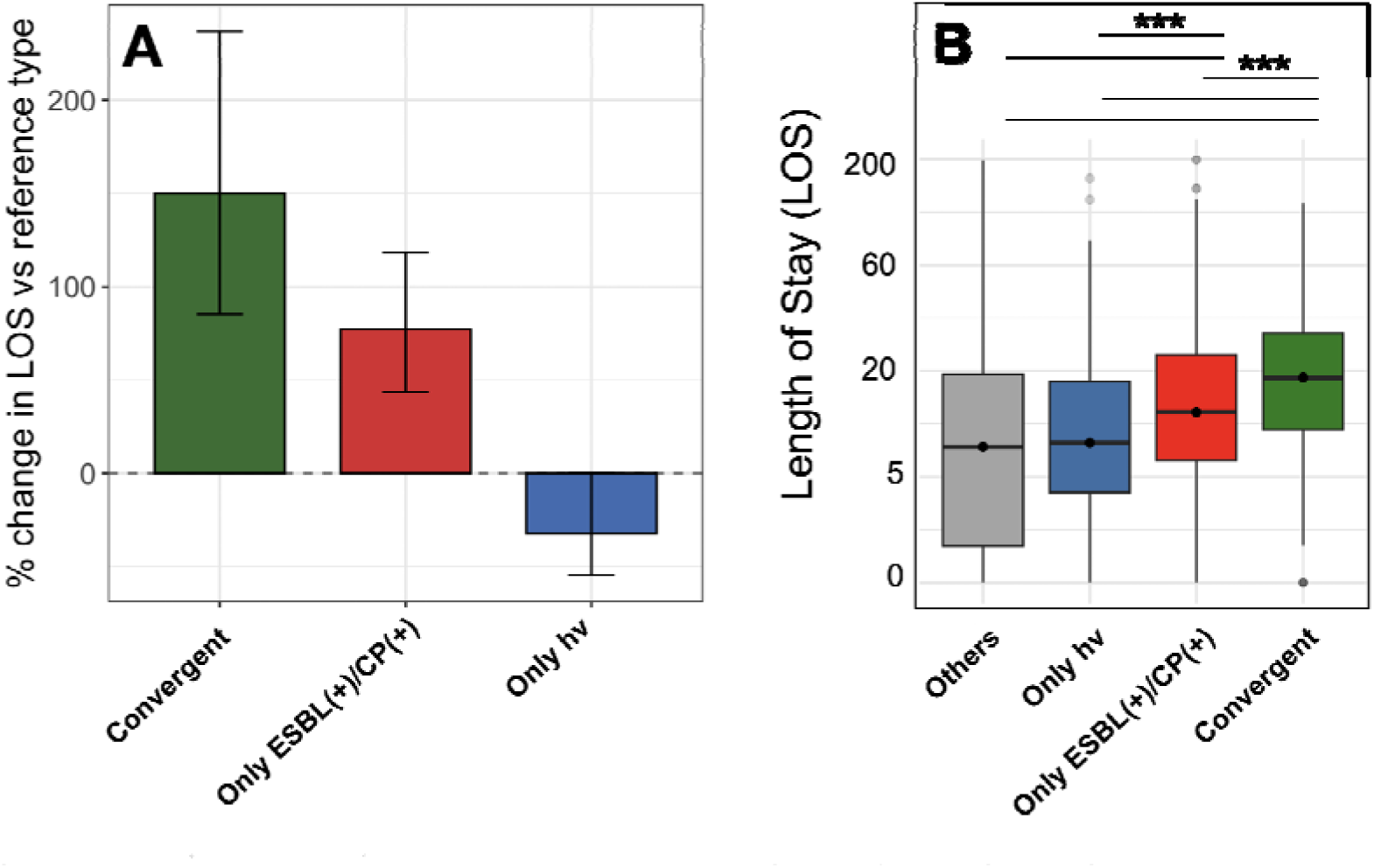
Association between virulence–resistance profiles and length of hospitalisation (LOS). A) Average change in hospitalization length for patients infected with *K. pneumoniae* isolates of differing virulence and resistance status. Values are inferred from a multivariable regression model accounting for clinical confounders (age, Charlson score, BMI, gender, infection source, and region). The reference category represents non-hypervirulent (non-hv) and ESBL(-)/CP(-) isolates. Error bars indicate 95% confidence intervals. B) Distribution of LOS among patients infected by isolates with distinct virulence–resistance profiles. Horizontal bar denote pairwise comparisons between subgroups, and asterisks indicate the significance level of differences between group means based on the Wilcoxon rank-sum test (*\*\*\** denotes *p* < 0.001).

### Predominant influence of antimicrobial resistance on infection severity

We dissected the genetic determinants of clinical severity by first estimating the associations between known virulence and resistance determinants and the three clinical outcome features (Model 1 in Methods) (Figure 6). After adjusting for confounding factors, nine known virulence/resistance determinants were found to be consistently linked with higher in-hospital mortality, ICU admission, and longer length of stay (LOS). These included the virulence factors *rmpA2*; colistin-resistance mutations in *mgrB and pmrB*; extended-spectrum β-lactamase production (*bla*_CTX-M-15_ gene); outer-membrane porin defects (OmpK36 TD mutations); and acquired carbapenemase genes — most notably the *bla*_OXA-48_ -like enzymes (*bla*_OXA-232_) (Figure 6). These determinants demonstrated a pleiotropic effect across multiple clinical phenotypes. Although individual genetic determinants may influence outcomes on their own, their co-occurrence on the same genomic background can lead to cross-resistance and obscure their individual effects. The gene-co-occurrence correlogram shows a tightly correlated cluster of hypervirulence genes, alongside a strong correlation among key multidrug-resistance determinants (colistin, carbapenem and cephalosporin resistance) (Figure 6). Hypervirulence genes exhibited moderate to strong but sporadic positive correlations with key resistance genes, including *bla*_OXA-48_-like carbapenemase genes, acquired ESBLs, colistin-resistance mutations and outer-membrane porin defects (OmpK35 deletion, OmpK36-TD mutations) (Figure 6). We also evaluated the effect of genetic co-occurrence on clinical severity by incorporating known virulence and resistance determinants alongside demographic and clinical covariates in a unified multivariate model (Model 2 in Methods) (Figure S3). The virulence genes showed no significant association with any of the outcomes. In contrast, various resistance-related categories demonstrated significant and consistent effects across all outcomes. Specifically, carbapenem and ESBL resistance determinants linked to markedly higher odds of in-hospital mortality and to substantially longer hospital stays (up to a 30-100 % increase in LOS) (Figure S3). Although resistance and virulence determinants frequently co-occurred within the same isolates, this overlap did not distort the effects of either factor in the regression models, as all indicators of collinearity between these genes remained within limits (Variance Inflation Factor (VIF) <5). Finally, the interaction model (Model 3) also demonstrated that higher resistance scores were independently associated with increased in-hospital mortality, higher ICU admission rates, and longer length of stay (pvalue < 0.05), whereas no significant effect of virulence level or resistance–virulence interaction was detected for any clinical outcome. Overall, these findings indicate that antimicrobial resistance traits, rather than hypervirulence alone, are the principal drivers of poor clinical outcomes, and that the co-occurrence of resistance and virulence further amplifies risk through additive effects rather than a synergistic interaction.

**Figure 6.**
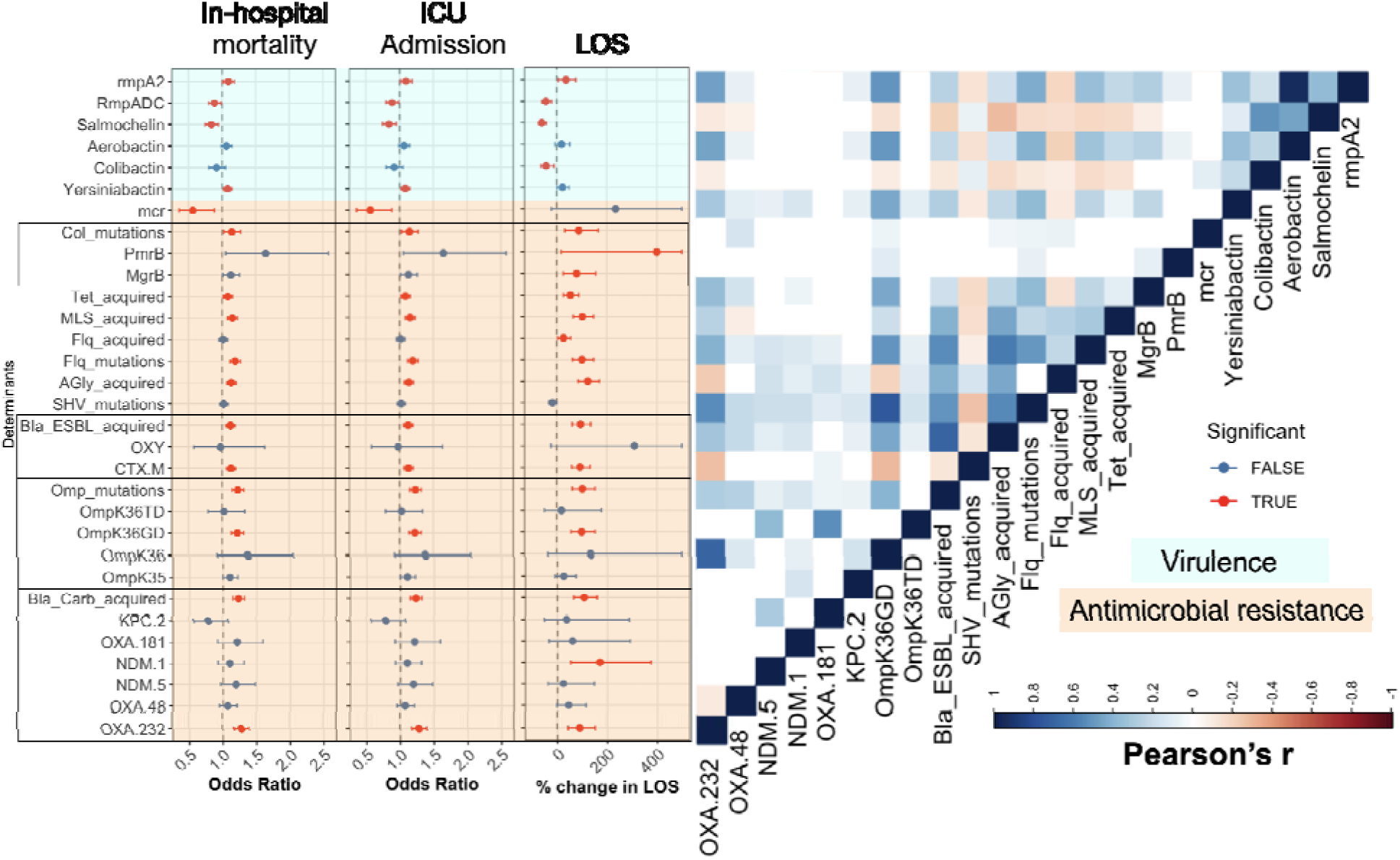
Association of virulence and resistance determinants with clinical outcomes. Odds ratios (ORs) are shown for in-hospital mortality and ICU (intensive care unit) admission, reflecting the strength of association between each genetic or clinical feature and these outcome after adjusting for clinical confounders. For LOS, the bars represent mean changes in LOS compared with the baseline group, i.e. exposure to isolates without the gene/mutation. Error bar indicate 95% confidence intervals (CIs) and are truncated for LOS to improve visual clarity. Rows correspond to key resistance and virulence categories: bla_Crab_acquired (carbapenemase genes), Omp_mutations (outer membrane porin mutation), and Col_mutations (colistin resistanc mutations in *mgrB* and *pmrB*). AGly, Flq, Tet, and MLS refer to resistance determinant (mutations or acquired genes) against aminoglycosides, fluoroquinolones, tetracyclines, and macrolide–lincosamide–streptogramin antimicrobials, respectively. Statistical significance (red lines) was assessed using Benjamini–Hochberg–corrected p values derived from the regression analyses. The correlogram displays pairwise co-occurance between features; non-significant correlations (pvalue > 0.05) are marked as white cells.

### Carbapenemase- and virulence-bearing plasmids are linked to increased clinical severity

The MDR and hv genes in *K. pneumoniae* co-occurrence and co-transfer are mediated via plasmid-borne elements, which serve as backbones for horizontal gene transfer of these genes. To determine which backbone confers a higher relative risk of clinical severity, we conducted a risk assessment of the plasmidome, which contained 179 plasmid fragments. The plasmidome is dominated by Col-type replicons, with frequent ColKP3–IncF co-carriage. Canonical backbone–enzyme associations include carbapenemase-encoding IncL/M– *bla*_OXA-48_-like, IncC/IncFII(K)–*bla*_NDM_, and ColKP3– *bla*_OXA-232_ plasmids, as well as large IncHI1B–IncF hybrids encoding both resistance and hypervirulence determinants (Figure S4). Despite variation across the associations with the three outcomes, carbapenemase-encoding plasmids, particularly those carrying *bla*_NDM-1_, *bla*_OXA-48_, and *bla*_OXA-232_ genes, were associated with an elevated risk of ICU admission and mortality, respectively. The IncL/M and ColKP3 plasmids, associated with *bla*_OXA-232_ and *bla*_OXA-48_, respectively, were previously shown to underlie carbapenem resistance in the high-risk dominant clone of ST2096 (Figure S4) [37, 40]. Plasmids containing the Col440I_1 replicon also exhibited a higher relative risk despite lacking major resistance determinants; this effect is likely due to their strong association with high-risk epidemic clones [41], indicating that Col440I_1 plasmid may serve as a predictive biomarker of strain-level virulence potential. The association with LOS showed a distinctive pattern in which large IncHI1B–IncF hybrid plasmids, which carried resistance genes, occasionally ESBL genes, and the *aerobactin* and *rmpA* gene cassette, exhibited a relatively stronger link with length of hospitalization (Figure S4). These plasmids have been shown to occur frequently in high-risk *K. pneumoniae* clones circulating regionally, including ST2096 and ST147 [42, 43]. These findings highlight that the replicon composition and gene cargo of plasmids shape distinct clinical trajectories, with specific backbone types being associated with adverse infection outcomes.

### bGWAS identifies known and potential resistance/virulence biomarkers for clinical severity outcomes

To pinpoint bacterial genetic determinants contributing to infection-severity outcomes beyond the known resistance and virulence loci, we performed a bacterial GWAS using accessory gene and core-genome SNPs as independent variables, and three binarised outcome variables (in-hospital mortality (discharged vs deceased), ICU admission (admitted vs not-admitted) and LOS (short vs prolonged hospitalisation)). After excluding hypothetical and mobile genetic element protein families, we identified 13, 34 and 45 gene families associated with mortality, ICU admission and LOS, respectively (Table S3). Of these, 14 genes showed pleiotropic associations with more than one outcome (Table S3). Mortality-associated gene families were clustered into seven co-occurrence groups that arose independently across multiple lineages (Figure S5, Table S3). Two groups comprised canonical virulence and resistance genes, namely the siderophore *ybt* locus (11 genes) and the ESBL gene *bla*_CTX-M-15_. Beyond these well-characterised determinants, we detected genes with experimentally supported roles in virulence, including Type VI secretion system (T6SS) components (*hcp/tssD*), which contribute to host-cell adhesion and invasion, as well as intra-microbial competition [44, 45]. Two additional groups contained loci with plausible mechanistic links to pathogenesis: metabolic-adaptation genes (i.e. *oadA, citT, ttdA/B, yvrE*), implicated in anaerobic growth and alternative carbon-source utilisation [46–48], and stress-tolerance/persistence loci (i.e. *dsbA, ssb, yfiB, higA/higB*) encoding systems that support protein folding under host-associated stress [49–52]. Although direct virulence roles have not been fully demonstrated for all of these genes, similar mechanisms have been proposed for genes with the same functions. The broader mortality GWAS identified a further 496 gene families, which included the aerobactin (*iuc*) genes and Type 1 fimbrial genes (*fimA/C/D*) that function as adhesins for host-cell attachment and infection establishment [53], together with multiple resistance genes such as *bla*_NDM-1_, *bla*_CTX-M-15_, *aac(6*′*)IIc/aac(6*′*)Ib*, *aadA2/aadA3*, *rmtF1*, *catB*, *dfrA21/dfrA12*, *arr/arr-2* and *ble*. For ICU admission and LOS analyses, we identified 131 and 357 gene families under a relaxed lineage-effect threshold, respectively. While the ICU-admission GWAS revealed only moderate associations with loci related to Type I fimbriae and aminoglycoside resistance genes, i.e. *aadA3/ant(3*″*)-Ia*, the LOS analysis recovered several of the same resistance and virulence biomarkers, including multiple resistance determinants, *ybt* loci, and Type VI secretion system proteins that were also associated with mortality, underscoring the pleiotropic effects of these genetic features across clinical outcomes (Figure S5).

### Core-genome SNP GWAS highlights transporter, envelope and metabolic loci as biomarkers of clinical severity

The GWAS pipeline on core-genome SNPs using stringent lineage-effect threshold identified 444, 33 and 729 variants associated with in-hospital mortality, ICU admission and LOS, respectively. Among these, 59, 4 and 32 were missense/stop-gain SNPs with likely functional effects (Table S4), and seven SNPs had pleiotropic effect and were associated with more than one outcome. The genes harbouring these SNPs span multiple functional categories, several with plausible roles in virulence. These include transporter and efflux systems, envelope-modification pathways and metabolic-adaptation mechanisms. In particular, the transporter category encompasses efflux pumps linked to drug resistance, ABC transporters, cell-envelope/lipid A–modification systems and sugar/ABC-transporter/RND-family efflux systems (*acrB/acrAB*), which in *K. pneumoniae* have been implicated in both resistance and virulence [54]. Additional implicated loci include metabolic-adaptation genes (2-oxo-3-deoxygalactonate kinase, iron-containing alcohol dehydrogenase, amidase/hydantoinase family), outer-membrane protein W (*ompW*), the envelope-associated protein PagP (involved in lipid A acylation) and the biofilm regulator BssR. Although variant-level associations between these genes and clinical outcomes have not been reported, their mechanistic plausibility in virulence, i.e. involvement in biofilm formation and envelope synthesis, supports their implication in host clinical severity [55].

### Integrated genomic and clinical modelling achieves high predictive accuracy for infection-severity outcomes

We evaluated the predictive information of bacterial genomic data for infection-severity outcomes. For all three outcomes, we compared baseline and ensemble models, using elastic net and gradient-boosted decision tree classifiers for in-hospital mortality and ICU admission, and a baseline quantile regressor and probabilistic ensemble regressor for LOS. The training data comprised clinical metadata alone and with progressively added genomic features, including antimicrobial resistance and virulence genes, accessory genes, SNPs and unitigs, as well as a setting with all genomic features only. As expected, model performance was always higher on the training data than on test data (Figure S6), while validation and test performance were closely aligned, indicating minimal overfitting and robust generalisation to unseen samples (Figure 7). When additional genomic features were included, the divergence between training and test performance increased, reflecting greater model complexity; however, this effect was consistently more moderate for ensemble models than for baseline models for both in-hospital mortality and ICU admission (Figure S6). For LOS prediction, the divergence between training and test performance was similar between ensemble and linear baseline models (Figure S6). Despite this divergence, the ensemble models achieved higher predictive accuracy overall, as evidenced by stronger correlations between predicted and observed LOS outcomes (Figure S7A). These results suggest that ensemble methods can better capture complex, nonlinear interaction structures in high-dimensional genomic-clinical data while controlling variance, yielding more generalisable and accurate predictions than simpler linear models that may fail to model such effects. Using the best-performing ensemble model, clinical data alone provided modest discrimination on test and validation sets (mean AUC 0.74 for mortality and 0.76 for ICU admission on test data), and progressively adding genomic features further improved performance to the maximum AUC values of 0.78 for mortality and 0.79 for ICU admission, respectively (Figure 7). For LOS prediction, models trained using clinical features alone achieved a correlation of 0.53 between observed and predicted values, which increased to 0.60 upon the inclusion of genomic predictors (Figure 7). Moreover, genomic features on their own carry substantial predictive information, achieving approximately 71% of the predictive performance achieved by the full clinical-plus-genomic models for LOS prediction. Similarly, models using genomic features alone retained 94 % and 93 % of the AUC achieved by the full clinical-plus-genomic models for mortality and ICU admission prediction, respectively (Figure 7). The best-performing ensemble models trained on combined clinical and genomic predictors, selected by maximal AUC for classification and maximal R^2^ for regression, demonstrated modest-to-strong performance on the independent test set. For mortality, the best model achieved an AUC of 0.78 (95% CI, 0.75–0.80), with an F1 score of 0.67 (0.63–0.70), precision of 0.67 (0.60–0.74), and recall of 0.67 (0.64–0.70). For ICU admission, the best model reached an AUC of 0.79 (95% CI, 0.75–0.83), with an F1 score of 0.83 (0.80–0.85), precision of 0.78 (0.76–0.80), and recall of 0.88 (0.85–0.91). For LOS, the best model yielded a correlation of 0.59 (95% CI, 0.55–0.63) between observed and predicted values (Figure 7). To facilitate future use, we deployed the unitig-based models as a standalone application (www.github.com/hjy1805/Kp_prediction). The application enables users to upload a genome assembly to predict clinical severity outcomes and to identify resistance- and virulence-associated predictive biomarkers within the input data.

**Figure 7.**
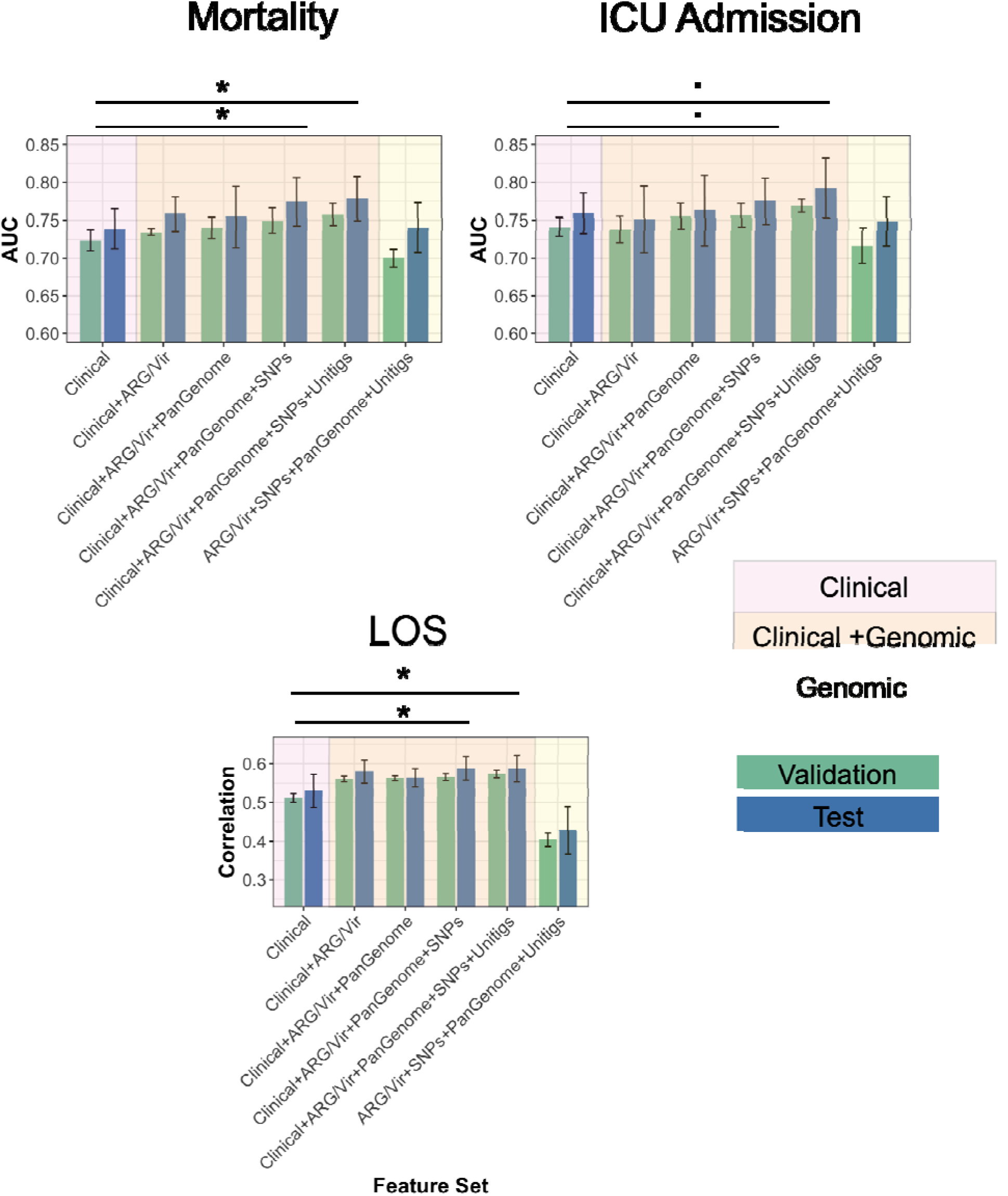
Performance of ensemble machine-learning models trained on clinical and genomic data. For in-hospital mortality and ICU admission, model performance was evaluated using the mean area under the receiver operating characteristic curve (AUC) obtained from tuned XGBoost classifiers. For LOS, performance was assessed using the correlation between observed and predicted values derived from NGBoost models on the independent test sets. Feature set included clinical variables, antimicrobial resistance genes (ARGs), virulence genes (Vir), pan-genome features, SNPs, and contig-based (unitig) features. Bars represent mean performance across five cross-validation folds, and error bars indicate the corresponding standard deviations. Symbols (·) and (*) denote one-sided Wilcoxon rank-sum test *p*-values of < 0.1 and < 0.05, respectively.

### Data efficiency, transferability, and uncertainty estimation of the models

Beyond high predictive accuracy, the trained models also exhibited properties considered essential for clinical decision-support, including robustness, generalisability, and quantification of predictive uncertainty. Model performance improved steadily with increasing training-data size but exhibited a diminishing-returns pattern beyond approximately 50% of the available dataset (Figure S8A). This trend indicates that the current cohort is sufficiently large to capture most of the predictive information present in the data, with only marginal gains expected from further expansion (Figure S8A). To assess the model’s transferability across healthcare settings, we also performed a leave-one-hospital-out validation: for each regional hospital, we held its data out as a test set and trained the model using data from all other hospitals, then evaluated performance on the left-out hospital’s test data (Figure S8B). The results demonstrate strong generalisability, with only a modest performance drop of 5% in AUC when applied to unseen regional data for two out of four regions. The probabilistic modelling also enabled estimation of prediction intervals for LOS, providing case-level measures of uncertainty around individual predictions. We evaluated the 90 % prediction interval coverage probability (PICP) for the models, which represents the proportion of observed LOS values that fell within the predicted range intended to contain future observations with 90 % probability (Figure S7B). Across models and input datasets, quantile regressors consistently outperformed NGBoost models, achieving higher PICP values that peaked at 85 %, indicating that the uncertainty estimates were well calibrated in predictive settings (Figure S7B). Moreover, the corresponding prediction interval width for the best-performing model was 1.07 times the standard deviation of the observed LOS (95 % CI: 0.96–1.20), indicating that the prediction uncertainty ranges remained relatively compact given the variability of the outcome. Taken together, these results suggest robust and well-calibrated uncertainty estimates with interval widths that are informative for clinical interpretation.

### Correlated clinical variables and resistance–virulence genetic determinants underlie model predictions

To pinpoint the most predictive genetic and clinical features, we used an interpretability approach that quantifies the contribution of each predictor to the model’s output, enabling ranking of feature importance. Using the model trained on clinical variables and virulence/resistance features, we identified the top predictors accounting for 90 % of the explained variance for in-hospital mortality (18 features), ICU admission (18 features) and LOS (20 features). Although the relative ranking of individual features varied slightly across outcomes, the most predictive variables were largely shared and exhibited correlated contributions across all three response variables (Pearson’s correlation *p* < 0.01) (Figure 8). Across outcomes, patient clinical data, including age, Charlson comorbidity index and body mass index, consistently ranked among the highest-impact predictors (Figure 8). For mortality prediction, fluoroquinolone-resistance mutations followed by the carbapenemase gene *bla*_OXA-232_ were the most predictive genetic features. Shared top genomic predictors across the outcomes included fluoroquinolone-resistance mutations, ESBLs, *bla*_OXA-232_, and porin gene mutations of OmpK36GD. In addition to strong correlations among clinical variables, the most predictive resistance and virulence genetic features, including carbapenemase genes, fluoroquinolone-resistance mutations and hypervirulence-associated siderophores, also exhibited high mutual correlation, reflecting co-occurrence of resistance and virulence determinants within the same genetic backgrounds (Figure 8). These indicate that while individual genetic determinants do not serve as dominant independent predictors of clinical severity relative to clinical features, they still provide prognostic information, and their interdependence with clinical factors yields multiple predictive biomarkers for infection outcomes. Given the strong correlations among the top predictors, we next assessed whether their effects were additive or involved interactions. We therefore examined nonlinear dependencies among predictors for all three outcomes and found the most consistent interactions among patient-level clinical variables, including age, Charlson comorbidity score, and infection source. These interactions were consistently negative, indicating that these variables capture partially redundant aspects of risk, such that their combined effect on outcome was weaker than the sum of their individual contributions (Figure S9). In contrast, only a small number of resistance–resistance interactions (fluoroquinolone and aminoglycoside determinants) and a single virulence–resistance interaction (yersiniabactin with fluoroquinolone resistance) showed modest but reproducible positive effects (Figure S9). Collectively, the sparse and low-magnitude interactions observed between genetic resistance and virulence features suggest that these predictors affect outcomes largely independently and additively, in agreement with our multivariate modeling results.

**Figure 8.**
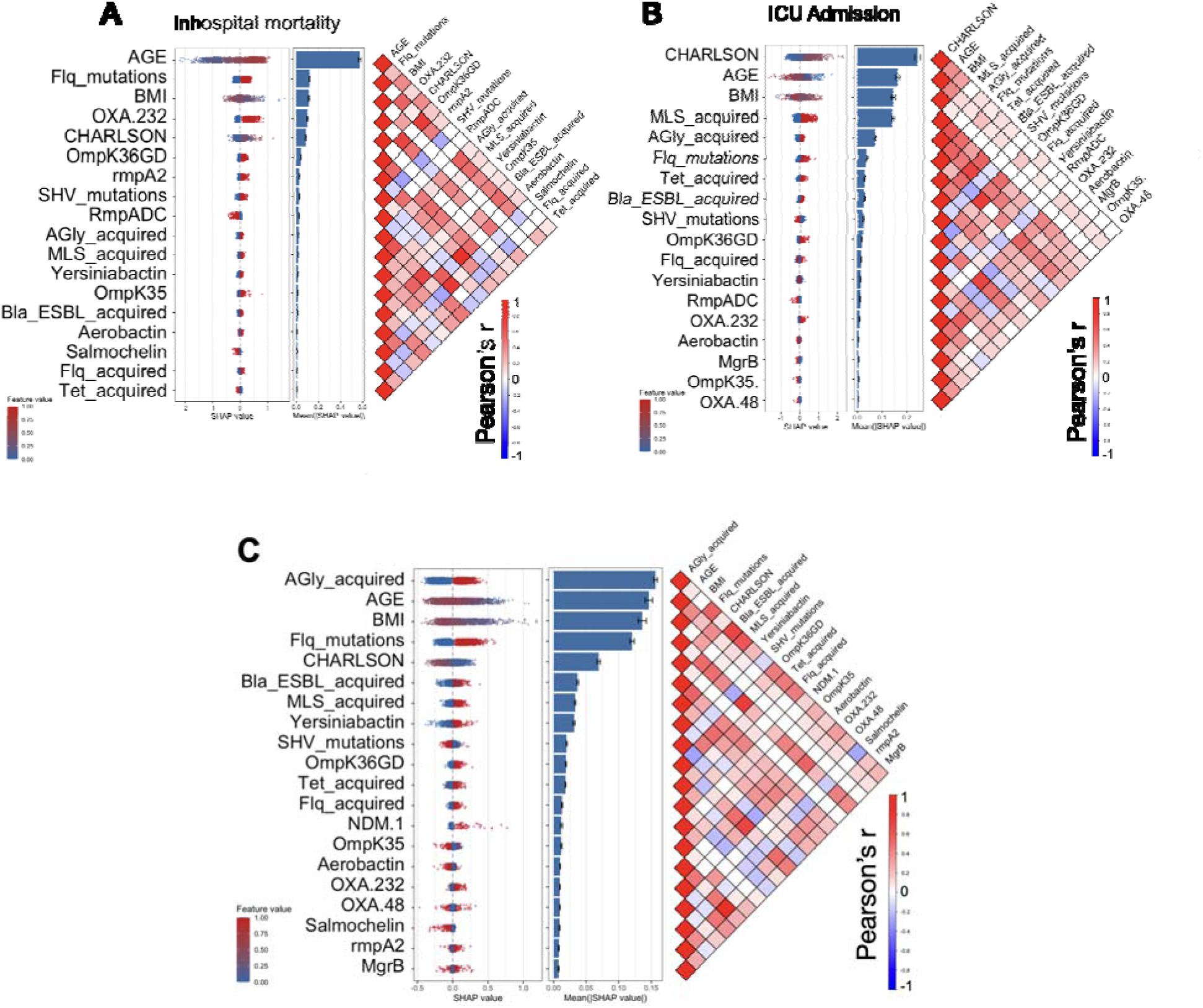
Feature importance analysis for (A) In-hospital mortality, (B) intensive care unit (ICU) admission, and (C) LOS prediction models. Each panel shows SHAP summary plots (left), displaying the relative contribution of individual features ranked by their mean absolute SHAP values, and Pearson correlation heat maps (right), illustrating inter-feature correlations among the top predictors that account for 90% of total feature contributions. In the correlograms, red and blue tones denote positive and negative Pearson correlations, respectively. Insignificant correlations are shown in white. Models were trained on input datasets containing clinical features together with pathogen resistance and virulence profiles. Abbreviations are the same a in Figure 6..

## Discussion

### Summary of results

We conducted a longitudinal, multicentre analysis of a large collection of *K. pneumoniae* isolates spanning seven years to dissect the clinical significance of circulating strains and the relationship between bacterial genomic variants, pathotypes and infection severity outcomes. By integrating multivariate regression, genome-wide association studies (GWAS), and machine-learning approaches, we identified genetic variants associated with in-hospital mortality, ICU admission, and LOS, including several previously unreported candidate markers of disease severity. We further examined the interplay between virulence and resistance determinants and their combined contribution to adverse clinical outcomes. Our analyses reveal that key hypervirulence and multidrug-resistance determinants are strongly associated with poor prognosis. In particular, isolates harbouring both resistance and virulence factors exhibited significantly higher mortality, increased ICU admission rates, and prolonged hospital stays. Carbapenemase genes emerged as some of the most powerful genomic predictors of clinical severity. These findings position ESBL(+)/CP(+)-hv *K. pneumoniae* as a critical public-health threat comparable to established healthcare-associated strains. Given the rising prevalence of dual-risk strains globally and the limited treatment options available for carbapenem-resistant infections, the development of genomic-based prognostic frameworks represents a vital step toward reducing mortality, morbidity, and the economic burden associated with CP *K. pneumoniae* infections.

### *K. pneumoniae* infections linked with convergent strains have poorer prognosis, and resistance remains the dominant statistical predictor of adverse outcomes

Previous studies of hospital-acquired *K. pneumoniae* infections have largely focused on mortality as the sole outcome and were often limited to single-centre cohorts, specific infection types (especially bloodstream infections), small sample sizes, or short study periods. In contrast, our study leverages a large, nationwide, diverse dataset spanning an extended timeframe and multiple clinical severity outcomes. In addition to offering new insights, our findings broadly align with earlier studies, despite some discrepancies. Our estimated mortality risk for ESBL(+)/CP(+)-only strains (19%) falls within the range of previous reports (10–20%) for these strains across different regions [56–61]. A recent survival analysis using a multivariable Cox regression model on a large-scale cohort of monomicrobial *K. pneumoniae* bloodstream infections in Norway over an 18-month period revealed an adjusted hazard ratio of 1.52 (CI 95% 0.98–2.38), which overlaps with our findings for ESBL(+)/CP(+) strains (1.46 [CI 95% 1.17–1.83]); however, that study did not specifically examine hazards for convergent strains [61]. Despite these broad congruencies, some differences were observed. In our study, the difference in LOS between CPKP and susceptible infections was substantial (median 46 vs. 14 days), exceeding that reported in a systematic review of single-hospital studies from China (approximately a ten-day difference) [62, 63]. Moreover, although patients infected with CPKP also had significantly higher odds of ICU admission compared with those without (OR: 2.25, 95% CI 1.46–3.55, p < 0.05), this effect size was smaller than the value reported in a previous single-hospital study [63]. In contrast to ESBL and carbapenemase status, hvKp alone was not associated with a significant increase in mortality or ICU admission rates in our cohort. A cohort study from China reported higher mortality among patients infected with hvKp compared with classical strains [20]. However, that study adopted a more stringent definition of hypervirulence, classifying hvKp as strains carrying all five virulence determinants (*rmpA/rmpA2, iuc, iro*, yersiniabactin, and hypermucoviscosity-associated genes). When applying the same strict definition, we observed a modestly increased odds ratio for mortality in hv strains compared with non-hv strains (OR: 1.74, 95% CI 0.83–3.36), although this did not reach statistical significance. The discrepancies between these findings and ours may reflect differences in sample composition and study design, as the previous analysis was restricted to a single-hospital, single-city setting, as well as variation in clinical management practices in different regions. Consistent with our findings, another study on hvKP strains did not find a statistically significant difference in mortality rates between hypervirulent and non-hypervirulent strains [64]. In the context of convergent strains, our results indicate a high degree of interdependence, when both resistance and virulence determinant predictors were included in the same model, only resistance remained significantly associated with clinical severity outcomes suggesting that resistance features capture most of the predictive variance linked to poor clinical outcomes. However, survival analysis demonstrated that infections caused by convergent strains had higher mortality than ESBL(+)/CP(+) infections, which is suggestive of additive biological effect that is partially masked by collinearity in multivariable models. This pattern indicates that resistance acts as the dominant statistical driver of mortality while virulence likely amplifies clinical severity within high-risk resistant lineages

### Integrative genomic approaches reveal multifactorial predictors of clinical severity

Our study applied bacterial GWAS and machine-learning approaches to infection outcomes rather than to the bacterium’s intrinsic traits such as resistance mechanisms or host specificity. Few studies have attempted to identify bacterial genomic biomarkers for the severity of host infections. These previous investigations, primarily in *Staphylococcus aureus* bacteraemia, assessed bacterial heritability for mortality and evaluated predictive performance, but reported only modest to weak heritability (< 5%) and limited predictive capacity [65, 66]. Other studies in *Streptococcus pneumoniae* and *Escherichia coli* likewise identified only weak associations between the infecting bacterial genome and major clinical endpoints, i.e. infective endocarditis, septic shock, ICU admission or mortality [67, 68]. In contrast, our results demonstrate a robust predictive power of bacterial genomic data for clinical severity in *K. pneumoniae* infections, thereby reinforcing the idea that pathoadaptation of *K. pneumoniae*, especially its virulence and resistance determinants, plays a key role in driving poor clinical outcomes. A prior study of carbapenem-resistant *K. pneumoniae* used machine learning models to distinguish colonization from infection and showed that both genomic and clinical data contribute to risk prediction (AUC ∼0.59-0.68 for combined models) [69]. Our study extends this by showing that other key clinical outcomes beyond colonization/infection status can also be predicted when machine learning is applied to comprehensive microbial genomic and clinical datasets.

### Combining GWAS and machine learning uncovers the full spectrum of virulence-and resistance-driven pathogenic mechanisms in natural strains

Beyond canonical resistance and virulence determinants, our GWAS also identified additional gene groups and SNPs that represent plausible and previously underexplored molecular targets linked to antimicrobial resistance/tolerance and pathogenicity. *K. pneumoniae* harbours an highly diverse genome, with more than half of its genes varying between isolates, enabling rapid adaptation through multiple virulence strategies [70, 71]. By integrating GWAS with machine-learning approaches, we were able not only to delineate the spectrum of variants associated with pathogenicity, but also to leverage their combined effects to predict infection severity. This framework complements the complex interplay between bacterial genetic variation, host factors, and treatment effects that together shape clinical outcomes. The moderate-to-strong performance of the predictive ensemble models indicates that the joint contribution of multiple genetic determinants captures information that is missed by conventional univariate models, including GWAS, which evaluate each variant in isolation. Importantly, although machine-learning models identify associations rather than causation, we show that strong genomic signatures—including carbapenemase and siderophore genes—serve as robust markers of poor clinical outcome. While these features may not directly drive disease severity, they nonetheless provide informative predictors. Given the tight interdependence between virulence and resistance determinants, combining multiple genomic biomarkers into diagnostic panels may enable improved prognostication of disease severity in *K. pneumoniae* infections, facilitating earlier risk stratification and more targeted therapeutic intervention.

### Limitations

Despite the scale and depth of this study, several limitations should be acknowledged. Our dataset is enriched for bloodstream isolates obtained from tertiary-care hospitals and therefore predominantly reflects lineages circulating within healthcare settings and associated with hospital-acquired infections. One other important limitation of our study is that community-associated strains are underrepresented in our dataset, so the full diversity of circulating populations may not be captured. This sampling bias may partly account for the lower apparent clinical severity of hvKp observed in our analyses. Although hvKp is well recognized for causing severe disease in otherwise healthy individuals in community settings, isolates collected through tertiary-care hospitals are more likely to be detected early and managed within structured clinical pathways, which can mitigate the severity of outcomes observed in hospital-based surveillance. Consequently, our findings may understate the true clinical impact of hvKp in community populations and emphasize the need for more inclusive sampling strategies in future studies. In addition, unlike many global collections that are dominated by classical hypervirulent or multidrug-resistant lineages such as ST11, ST258, ST307, ST15 and ST147 [72], our cohort is characterized by a high prevalence of a regionally high-risk clone of ST2096. This distinctive sequence type distribution indicates that, although broad biological trends are conserved, the local genomic landscape is skewed. Consequently, the estimated effect sizes and specific associations reported here may not be directly generalisable to other geographical or epidemiological contexts.

### Conclusion

Our results show that bacterial genomic data, when integrated with clinical metadata, can predict mortality and ICU admission with high accuracy, outperforming models based on clinical features alone. This capability reframes the management of *K. pneumoniae* infections: rather than relying solely on clinical deterioration signs, genomic biomarkers can be used at the time of diagnosis to identify patients at elevated risk and inform earlier, more intensive management. With whole-genome sequencing now achievable within 24–48 h, high-risk genomic signatures associated with severe outcomes can be identified during the initial phase of care. This information has the potential to support timely escalation of antimicrobial therapy, optimization of infection-control measures, and more strategic allocation of limited healthcare resources. At a population level, our framework provides a foundation for integrated genomic–clinical surveillance, enabling the rapid identification of emerging high-risk clones and coordinated responses across healthcare systems. Future efforts should focus on embedding genomic risk prediction into routine microbiology workflows, prospectively evaluating the clinical impact of genomics-guided management, and establishing international collaborations for global standardized genomic–clinical surveillance. Together, these steps could support a transition from predominantly reactive management toward a more proactive, data-driven approach to *K. pneumoniae* infections.

## Acknowledgements

The authors gratefully acknowledge the KAUST Bioscience Core Lab facility and staff for their support & assistance in this work.

## Data Accession

All short- and long-read sequencing data have been submitted to the European Nucleotide Archive (ENA) under accession number PRJEB36683. Assembly files have been deposited in GenBank under accession number PRJNA1116866. Metadata are provided in Supplementary Table S1 and in the GitHub directory.

## Authors contribution

RYA, MB, AP, and DM designed the study. RYA, MB, AP, and DM supervised the project and acquired funding. MMa, RYA, JH, OF, MM, and GZ conducted the research and generated the primary datasets. JH and DM developed and implemented the machine-learning platform. RH, SI, MBan, YL, AS, SMH, NB, DA, ASA, AWA, SMA, MMK, BA, MA, MEA, HSS, and SZ contributed resources, clinical materials, or analytical support. RYA, MM, AS, MB, AP, and DM wrote and edited the manuscript.

## Conflict of ineterst declaration

Authors declare no conflict of ineterst

## Notes

### Competing Interest Statement

The authors have declared no competing interest.

### Funding Statement

This study was supported by the KAUST Faculty Baseline Fund (BAS/1/1108-01-01) and the KAUST Baseline Fund (BAS/1/1020-01-01).

### Author Declarations

This study was approved by Institutional Review Board (IRB) of King Abdullah University for Science and Technology (approval number 17IBEC38) and from the Saudi Ministry of National Guard Health Affairs (MNGHA) (approval number NRC23R/533/08). The research described here was conducted in accordance with the Helsinki declaration.

## Reference

1. Paczosa, M.K. and J. Mecsas, Klebsiella pneumoniae: Going on the Offense with a Strong Defense. Microbiol Mol Biol Rev, 2016. 80(3): p. 629–61.

2. Lou, T., et al., Risk factors for infection and mortality caused by carbapenem-resistant Klebsiella pneumoniae: A large multicentre case-control and cohort study. J Infect, 2022. 84(5): p. 637–647.

3. Li, J., et al., Rapid emergence, transmission, and evolution of KPC and NDM coproducing carbapenem-resistant Klebsiella pneumoniae. Microbiol Res, 2025. 293: p. 128049.

4. Qiu, M., et al., Clinical Characterization, Risk Factors, and Mortality in Patients with Carbapenem-Resistant Hypervirulent Klebsiella pneumoniae Intra-Abdominal Infections. Infect Drug Resist, 2025. 18: p. 3647–3660.

5. Cao, Z., et al., Risk Factors for a Hospital-Acquired Carbapenem-Resistant Klebsiella pneumoniae Bloodstream Infection: A Five-Year Retrospective Study. Infect Drug Resist, 2022. 15: p. 641–654.

6. Plachouras, D., et al., Antimicrobial use in European acute care hospitals: results from the second point prevalence survey (PPS) of healthcare-associated infections and antimicrobial use, 2016 to 2017. Euro Surveill, 2018. 23(46).

7. Zhang, J., et al., Economic burden of carbapenem-resistant Klebsiella pneumoniae infections in Chinese hospitals: A 2019 analysis. J Glob Antimicrob Resist, 2025. 46: p. 63–70.

8. David, S., et al., Epidemic of carbapenem-resistant Klebsiella pneumoniae in Europe is driven by nosocomial spread. Nat Microbiol, 2019. 4(11): p. 1919–1929.

9. Zong, Z., A. Wu, and B. Hu, Infection Control in the Era of Antimicrobial Resistance in China: Progress, Challenges, and Opportunities. Clin Infect Dis, 2020. 71(Suppl 4): p. S372–S378.

10. Lin, X.C., et al., The Global and Regional Prevalence of Hospital-Acquired Carbapenem-Resistant Klebsiella pneumoniae Infection: A Systematic Review and Meta-analysis. Open Forum Infect Dis, 2024. 11(2): p. ofad649.

11. Russo, T.A. and C.M. Marr, Hypervirulent Klebsiella pneumoniae. Clin Microbiol Rev, 2019. 32(3).

12. Wei, D.D., et al., Emergence of KPC-producing Klebsiella pneumoniae hypervirulent clone of capsular serotype K1 that belongs to sequence type 11 in Mainland China. Diagn Microbiol Infect Dis, 2016. 85(2): p. 192–4.

13. Zhang, R., et al., Emergence of Carbapenem-Resistant Serotype K1 Hypervirulent Klebsiella pneumoniae Strains in China. Antimicrob Agents Chemother, 2016. 60(1): p. 709–11.

14. Chen, J., et al., Characteristics and risk factors for infection and mortality caused by Klebsiella pneumoniae in patients with acute pancreatitis. Front Public Health, 2024. 12: p. 1533765.

15. Jia, Y., et al., Clinical Characteristics, Drug Resistance, and Risk Factors for Death of Klebsiella pneumoniae Infection in Patients with Acute Pancreatitis: A Single-Center Retrospective Study from China. Infect Drug Resist, 2023. 16: p. 5039–5053.

16. Wu, C., L. Zheng, and J. Yao, Analysis of Risk Factors and Mortality of Patients with Carbapenem-Resistant Klebsiella pneumoniae Infection. Infect Drug Resist, 2022. 15: p. 2383–2391.

17. Xu, L., X. Sun, and X. Ma, Systematic review and meta-analysis of mortality of patients infected with carbapenem-resistant Klebsiella pneumoniae. Ann Clin Microbiol Antimicrob, 2017. 16(1): p. 18.

18. Huang, W., et al., In-hospital Medical Costs of Infections Caused by Carbapenem-resistant Klebsiella pneumoniae. Clin Infect Dis, 2018. 67(suppl_2): p. S225–S230.

19. Zhou, R., et al., Impact of carbapenem resistance on mortality in patients infected with Enterobacteriaceae: a systematic review and meta-analysis. BMJ Open, 2021. 11(12): p. e054971.

20. Tang, Y., et al., Genomically defined hypervirulent Klebsiella pneumoniae contributed to early-onset increased mortality. Nat Commun, 2025. 16(1): p. 2096.

21. Zhao, Y., et al., An Outbreak of Carbapenem-Resistant and Hypervirulent Klebsiella pneumoniae in an Intensive Care Unit of a Major Teaching Hospital in Wenzhou, China. Front Public Health, 2019. 7: p. 229.

22. Gu, D., et al., A fatal outbreak of ST11 carbapenem-resistant hypervirulent Klebsiella pneumoniae in a Chinese hospital: a molecular epidemiological study. Lancet Infect Dis, 2018. 18(1): p. 37–46.

23. Wick, R.R., et al., Unicycler: Resolving bacterial genome assemblies from short and long sequencing reads. PLoS Comput Biol, 2017. 13(6): p. e1005595.

24. Schwengers, O., et al., Bakta: rapid and standardized annotation of bacterial genomes via alignment-free sequence identification. Microb Genom, 2021. 7(11).

25. Tonkin-Hill, G., et al., Producing polished prokaryotic pangenomes with the Panaroo pipeline. Genome Biol, 2020. 21(1): p. 180.

26. Lam, M.M.C., et al., A genomic surveillance framework and genotyping tool for Klebsiella pneumoniae and its related species complex. Nat Commun, 2021. 12(1): p. 4188.

27. Hoashi, K., et al., Community-acquired liver abscess caused by capsular genotype K2-ST375 hypervirulent Klebsiella pneumoniae isolates. IDCases, 2019. 17: p. e00577.

28. Paradis, E., J. Claude, and K. Strimmer, APE: Analyses of Phylogenetics and Evolution in R language. Bioinformatics, 2004. 20(2): p. 289–90.

29. James G, W.D., Hastie T, Tibshirani R, An Introduction to Statistical Learning. 2017: Springer.

30. Wick, R.R., et al., Bandage: interactive visualization of de novo genome assemblies. Bioinformatics, 2015. 31(20): p. 3350–2.

31. Brynildsrud, O., et al., Rapid scoring of genes in microbial pan-genome-wide association studies with Scoary. Genome Biol, 2016. 17(1): p. 238.

32. Holley, G. and P. Melsted, Bifrost: highly parallel construction and indexing of colored and compacted de Bruijn graphs. Genome Biol, 2020. 21(1): p. 249.

33. Ester, M., H.P. Kriegel, and X. Xu, XGBoost: A scalable tree boosting system. In Proceedings of the 22Nd ACM SIGKDD International Conference on Knowledge Discovery and Data Mining *(vol , pg* 785, *2016).* Geographical Analysis, 2022.

34. Duan, T., et al., NGBoost: Natural Gradient Boosting for Probabilistic Prediction. International Conference on Machine Learning, Vol 119, 2020. **119**.

35. Mitchell, R., et al., Sampling Permutations for Shapley Value Estimation. Journal of Machine Learning Research, 2022. 23: p. 1–46.

36. Hala, S., et al., The emergence of highly resistant and hypervirulent Klebsiella pneumoniae CC14 clone in a tertiary hospital over 8 years. Genome Med, 2024. 16(1): p. 58.

37. Huang, J., et al., The dissemination of multidrug-resistant and hypervirulent Klebsiella pneumoniae clones across the Kingdom of Saudi Arabia. Emerg Microbes Infect, 2024. 13(1): p. 2427793.

38. Zhang, Y., et al., Sequence-Based Genomic Analysis Reveals Transmission of Antibiotic Resistance and Virulence among Carbapenemase-Producing Klebsiella pneumoniae Strains. mSphere, 2022. 7(3): p. e0014322.

39. Luterbach, C.L., et al., Transmission of Carbapenem-Resistant Klebsiella pneumoniae in US Hospitals. Clin Infect Dis, 2023. 76(2): p. 229–237.

40. David, S., et al., Integrated chromosomal and plasmid sequence analyses reveal diverse modes of carbapenemase gene spread among Klebsiella pneumoniae. Proc Natl Acad Sci U S A, 2020. 117(40): p. 25043–25054.

41. Medina, V.A., et al., Epidemiological Genomics of Klebsiella pneumoniae isolates from hospitals across Colombia. NPJ Antimicrob Resist, 2025. 3(1): p. 64.

42. Shankar, C., et al., Hybrid Plasmids Encoding Antimicrobial Resistance and Virulence Traits Among Hypervirulent Klebsiella pneumoniae ST2096 in India. Front Cell Infect Microbiol, 2022. 12: p. 875116.

43. Turton, J.F., et al., Klebsiella pneumoniae sequence type 147: a high-risk clone increasingly associated with plasmids carrying both resistance and virulence elements. J Med Microbiol, 2024. 73(4).

44. Bray, A.S., et al., Klebsiella pneumoniae employs a type VI secretion system to overcome microbiota-mediated colonization resistance. Nat Commun, 2025. 16(1): p. 940.

45. Hsieh, P.F., et al., Klebsiella pneumoniae Type VI Secretion System Contributes to Bacterial Competition, Cell Invasion, Type-1 Fimbriae Expression, and In Vivo Colonization. J Infect Dis, 2019. 219(4): p. 637–647.

46. Chen, Y.T., et al., Genomic diversity of citrate fermentation in Klebsiella pneumoniae. BMC Microbiol, 2009. 9: p. 168.

47. Liu, M., et al., CitAB Two-Component System-Regulated Citrate Utilization Contributes to Vibrio cholerae Competitiveness with the Gut Microbiota. Infect Immun, 2019. 87(3).

48. Bachman, M.A., et al., Genome-Wide Identification of Klebsiella pneumoniae Fitness Genes during Lung Infection. mBio, 2015. 6(3): p. e00775.

49. Eger, E., et al., Extensively Drug-Resistant Klebsiella pneumoniae Counteracts Fitness and Virulence Costs That Accompanied Ceftazidime-Avibactam Resistance Acquisition. Microbiol Spectr, 2022. 10(3): p. e0014822.

50. Huang, Y.H. and C.Y. Huang, Characterization of a single-stranded DNA-binding protein from Klebsiella pneumoniae: mutation at either Arg73 or Ser76 causes a less cooperative complex on DNA. Genes Cells, 2012. 17(2): p. 146–57.

51. Huertas, M.G., et al., Klebsiella pneumoniae yfiRNB operon affects biofilm formation, polysaccharide production and drug susceptibility. Microbiology (Reading), 2014. 160(Pt 12): p. 2595–2606.

52. Qi, Q., M. Kamruzzaman, and J.R. Iredell, The higBA-Type Toxin-Antitoxin System in IncC Plasmids Is a Mobilizable Ciprofloxacin-Inducible System. mSphere, 2021. 6(3): p. e0042421.

53. Struve, C., M. Bojer, and K.A. Krogfelt, Identification of a conserved chromosomal region encoding Klebsiella pneumoniae type 1 and type 3 fimbriae and assessment of the role of fimbriae in pathogenicity. Infect Immun, 2009. 77(11): p. 5016–24.

54. Padilla, E., et al., Klebsiella pneumoniae AcrAB efflux pump contributes to antimicrobial resistance and virulence. Antimicrob Agents Chemother, 2010. 54(1): p. 177–83.

55. Nguyen, T.N.T., G. Howells, and F.L. Short, How Klebsiella pneumoniae controls its virulence. PLoS Pathog, 2025. 21(9): p. e1013499.

56. Meatherall, B.L., et al., Incidence, risk factors, and outcomes of Klebsiella pneumoniae bacteremia. Am J Med, 2009. 122(9): p. 866–73.

57. Kim, D., et al., Antimicrobial resistance and virulence factors of Klebsiella pneumoniae affecting 30 day mortality in patients with bloodstream infection. J Antimicrob Chemother, 2019. 74(1): p. 190–199.

58. Maatallah, M., et al., Klebsiella variicola is a frequent cause of bloodstream infection in the stockholm area, and associated with higher mortality compared to K. pneumoniae. PLoS One, 2014. 9(11): p. e113539.

59. Imai, K., et al., Clinical characteristics in blood stream infections caused by Klebsiella pneumoniae, Klebsiella variicola, and Klebsiella quasipneumoniae: a comparative study, Japan, 2014-2017. BMC Infect Dis, 2019. 19(1): p. 946.

60. Brady, M., et al., Klebsiella pneumoniae bloodstream infection, antimicrobial resistance and consumption trends in Ireland: 2008 to 2013. Eur J Clin Microbiol Infect Dis, 2016. 35(11): p. 1777–1785.

61. Fostervold, A., et al., Risk of death in Klebsiella pneumoniae bloodstream infections is associated with specific phylogenetic lineages. J Infect, 2024. 88(5): p. 106155.

62. Yao, Y., et al., Healthcare-associated carbapenem-resistant Klebsiella pneumoniae infections are associated with higher mortality compared to carbapenem-susceptible K. pneumoniae infections in the intensive care unit: a retrospective cohort study. J Hosp Infect, 2024. 148: p. 30–38.

63. Li, Y., et al., Five-year change of prevalence and risk factors for infection and mortality of carbapenem-resistant Klebsiella pneumoniae bloodstream infection in a tertiary hospital in North China. Antimicrob Resist Infect Control, 2020. 9(1): p. 79.

64. Hwang, J.H., et al., Clinical Features and Risk Factors Associated With 30-Day Mortality in Patients With Pneumonia Caused by Hypervirulent Klebsiella pneumoniae (hvKP). Ann Lab Med, 2020. 40(6): p. 481–487.

65. Giulieri, S.G., et al., A statistical genomics framework to trace bacterial genomic predictors of clinical outcomes in Staphylococcus aureus bacteremia. Cell Rep, 2023. 42(9): p. 113069.

66. Recker, M., et al., Clonal differences in Staphylococcus aureus bacteraemia-associated mortality. Nat Microbiol, 2017. 2(10): p. 1381–1388.

67. Denamur, E., et al., Genome wide association study of Escherichia coli bloodstream infection isolates identifies genetic determinants for the portal of entry but not fatal outcome. PLoS Genet, 2022. 18(3): p. e1010112.

68. Lees, J.A., et al., Joint sequencing of human and pathogen genomes reveals the genetics of pneumococcal meningitis. Nat Commun, 2019. 10(1): p. 2176.

69. Lapp, Z., et al., Patient and Microbial Genomic Factors Associated with Carbapenem-Resistant Klebsiella pneumoniae Extraintestinal Colonization and Infection. mSystems, 2021. 6(2).

70. Holt, K.E., et al., Genomic analysis of diversity, population structure, virulence, and antimicrobial resistance in Klebsiella pneumoniae, an urgent threat to public health. Proc Natl Acad Sci U S A, 2015. 112(27): p. E3574–81.

71. Martin, R.M. and M.A. Bachman, Colonization, Infection, and the Accessory Genome of Klebsiella pneumoniae. Front Cell Infect Microbiol, 2018. 8: p. 4.

72. Heng, H., et al., Global genomic profiling of Klebsiella pneumoniae: A spatio-temporal population structure analysis. Int J Antimicrob Agents, 2024. 63(2): p. 107055.

73. Gasparini, A., comorbidity: An R package for computing comorbidity scores. Journal of Open Source Software, 2018.

74. Jombart, T. and I. Ahmed, adegenet 1.3-1: new tools for the analysis of genome-wide SNP data. Bioinformatics, 2011. 27(21): p. 3070–1.

75. Croucher, N.J., et al., Rapid phylogenetic analysis of large samples of recombinant bacterial whole genome sequences using Gubbins. Nucleic Acids Res, 2015. 43(3): p. e15.

