## Supplementary Materials for "Genomic Signatures and Prediction of Clinical Severity in *Klebsiella pneumoniae* infections in a Multicenter Cohort"

### Supplementary Methods/Figures/Tables

#### Supplementary Methods

##### Ethical approvals

This study was approved by Institutional Review Board (IRB) of King Abdullah University for Science and Technology (approval number 17IBEC38) and from the Saudi Ministry of National Guard Health Affairs (MNGHA) (approval number NRC23R/533/08). The research described here was conducted in accordance with the Helsinki declaration.

##### Sample collection and description

Between 2018 and 2025, a total of 1,178 non-duplicate clinical isolates of the *Klebsiella pneumoniae* complex were collected from 1,085 patients diagnosed with infection as part of a multicenter hospital surveillance program. During this period the participating hospitals reported 48,125 cases of *K. pneumoniae* complex infections, meaning our collection represents approximately 2% of all *K. pneumoniae* complex infections. The isolates originated from tertiary-care facilities of the Ministry of National Guard Health Affairs (MNGHA) located in Riyadh, Jeddah, Madinah, Al-Ahsa and Dammam. The 1,178 isolates originated from hospitals in the following cities/regions: Riyadh (central region) (n = 593), Al-Ahsa (eastern region) (n = 190), Dammam (eastern region) (n = 86), Madinah (north western region) (n = 167), and Jeddah (western region) (n = 142). We included a subset from the MNGHA hospital in Jeddah: 128 isolates previously sequenced (2018–2022) from an earlier genomic study [36]. These were re-analysed to maintain consistent annotation and comparability across all sites.

Clinical samples were obtained from diverse infection sites, with the majority derived from bloodstream infections, followed by respiratory tract and urinary tract infections; additional isolates were sampled from surgical-site wounds, other wound infections, and sterile body fluids including ascitic fluid, pleural fluid and ocular specimens. Sampling was performed within routine diagnostic workflows in hospital microbiology laboratories, and isolates were identified to the species level using standard microbiological procedures prior to genomic analysis. Comprehensive patient-level metadata—including demographic, clinical, and sample-related information—were systematically extracted from electronic health records (EHRs). Demographic variables comprised patient age, gender, nationality and body mass index (BMI). Clinical details included dates of admission, discharged (or deceased), length of hospital stay (LOS), ward or unit of admission (adult, pediatric, ICU or specialty ward), clinical diagnosis and comorbidity status. Community-acquired infections are defined as infections detected ≤ 48 hours of hospital admission; infections detected > 48 hours are classified as nosocomial. Comorbidity scores were computed as the Charlson score using ICD-10 codes and the R package comorbidity [73]. The majority of isolates were *K. pneumoniae sensu stricto* (1,115; 94.7 %) followed by *K. quasipneumoniae* (42; 3.6 %), *K. variicola* (11; 0.9 %), K. michiganensis* (4; 0.3 %), *K. aerogenes* (4; 0.2 %) and *K. pasteurii* (2; 0.2 %). The specifications of the samples are provided in Supplemental Tables S1 and S2.

##### Antimicrobial susceptibility testing

Following initial isolation on MacConkey agar, bacterial colonies were subjected to identification and antimicrobial susceptibility testing (AST) using the DxM MicroScan WalkAway ID/AST System (Beckman Coulter). Well-isolated colonies were suspended in sterile saline, and the inoculum was standardized to a 0.5 McFarland turbidity equivalent using the Prompt Inoculation System to ensure uniform cell density. Standardized suspensions were dispensed into MicroScan seed trays and transferred into dehydrated ID/AST panels using the Renok inoculator, thereby rehydrating biochemical substrates and serial antimicrobial dilutions contained within the wells. The panels were then sealed, barcoded, and placed into the automated WalkAway instrument, where incubation at 35–37 °C was carried out for 16–24 hours under controlled conditions. During incubation, bacterial growth in each well was continuously assessed by the instrument's photometric and fluorometric detection systems. Growth inhibition patterns were analyzed by the LabPro software, which determined the minimum inhibitory concentration (MIC) for each antimicrobial tested and automatically assigned susceptibility categories (susceptible, intermediate, resistant) according to the European Committee on Antimicrobial Susceptibility Testing (EUCAST) breakpoints. For quality control, *K. quasipneumoniae* subsp. *similipneumoniae* ATCC 700603 was included in each batch of tests, alongside internal system controls, to verify panel performance and ensure validity of results.

**Short read whole genome sequencing**

Samples were genome extracted using the Quiagnen DNAeasy Blood &Tissue kit, following instructions for Gram negative bacteria. Whole-genome sequencing libraries were prepared using the BGI optimal DNA Library Prep Kit (BGI, Shenzhen, China). Briefly, genomic DNA was enzymatically fragmented and subjected to magnetic bead-based size selection to obtain fragments of appropriate length. The selected fragments were end-repaired to generate blunt ends, followed by A-tailing and ligation of sequencing adapters. The adapter-ligated products were enriched by limited-cycle PCR amplification and assessed for quality and yield prior to sequencing. Prepared double-stranded libraries were denatured into single-stranded templates, which were circularized to form single-stranded circular DNA molecules. Residual linear DNA fragments were removed enzymatically. The circularized templates were then subjected to rolling circle amplification (RCA) with phi29 polymerase, generating highly uniform DNA NanoBalls (DNBs). Each DNB carried approximately 300 copies of the original single-stranded template, thereby enhancing signal strength and sequencing accuracy. DNBs were loaded onto patterned nanoarrays and sequenced on the DNBSEQ-G400 platform (BGI, Shenzhen, China). Sequencing was performed in paired-end mode with a read length of 150 base pairs (PE150).

**Epidemiological cluster analysis**

To identify epidemiological clusters in the collection across all lineages, we employed a hybrid k-mer and alignment-based approach. First, we enumerated 50-bp k-mers in the collection and calculated the pairwise number of k-mers shared between genomes. For genomes from the same patients, we considered the genome with the oldest isolation date to represent that patient's strain in the clustering analysis, thereby avoiding bias from multiple samples of the same infection when defining epidemiological clusters. We then computed the Jaccard distance from this k-mer sharing matrix. We applied the R package adegenet (v1.3-1) on the Jaccard distance matrix [74] to infer maximum-likelihood networks based on pairwise Jaccard distances between samples, taking into account their isolation dates. To define transmission clusters, we used the similarity kmer-based distance cut-off that corresponded to 20 SNPs, a threshold reported for detecting transmission events in cross-country hospital settings for *K pneumoniae* [8]. To identify the equivalent cut-off value to 20 SNPs, we mapped the short-reads to a local reference genome of ST2096. This reference was constructed by concatenating the contigs of the isolate with the best assembly statistics (highest N50). We then removed hypervariable sites using Gubbins (v.3.3.1) [75] and computed pairwise SNP distances using distdna in the ape package. We then determined the mean Jaccard distance among all genomes with pairwise SNP distances ≤ 20 and used this as the cut-off in the k-mer-based analysis to identify epidemiological clusters. The resulting network and modules were extracted, analyzed, and visualized using the igraph library (v1.5.1) in R. We used the package Epitools package (v0.5.10.1) to estimate the association of an isolate being part of a cluster and the genetic or host clinical status.

#### Supplementary Tables/Figures

**Supplementary Table S1:** Accession number and associated metadata for the isolates, included in the study, provided as SupplumentalTable1.csv.

**Supplementary Table S2.** Characteristics of the isolates and the patients from whom the isolates were retrieved.

* Include ischemic heart disease, heart failure, and stroke history.
†Includes COPD and asthma.
‡Includes dementia, seizure disorders, and neuromuscular disease.

| Patients (n = 1,085) | |
|---|---|
| **Gender** | **n (%)** |
| Male | 537 (49.5) |
| Female | 548 (50.5) |
| Age (Interquartile Range) | 61 (38–74) |
| **Diagnosis** | **n (%)** |
| Leukemia | 26 (2.3) |
| Urinary tract infection | 215 (19.8) |
| Sepsis | 115 (10.6) |
| Septic Shock | 62 (5.7) |
| Pneumonia | 175 (16.1) |
| **Comorbidity** | **n (%)** |
| Diabetes Mellites | 267 (24.6) |
| Hypertension | 166 (15.3) |
| Chronic lung disease† | 32 (2.9) |
| Malignancy | 104 (9.6) |
| Cardiovascular disease* | 121 (11.2) |
| Chronic kidney disease | 120 (11.0) |
| Neurological disorders‡ | 114 (10.5) |
| Liver disease | 22 (2.0) |
| **Overall mortality** | **n (%)** |
| Died | 313 (30.9) |
| Alive | 831 (69.1) |
| **Mortality by diagnosis and care settings** | **n (%)** |
| ICU/NICU | 80/104 (76.9) |
| Other wards | 233/977 (23.8) |
| Sepsis | 44/115 (38.3) |
| Septic shock | 39/62 (62.9) |
| Pneumonia | 73/175 (41.7) |
| Urinary tract infection | 26/215 (12.1) |
| Leukemia | 9/26 (34.6) |
| Other diagnosis | 163/592 (27.5) |

| Length of Stay (LOS) by infection type | Days – median (IQR) |
|---|---|
| Community-acquired | 10 (5-18) |
| Hospital-acquired | 54 (24-112) |

| Isolates (n = 1,178) | |
|---|---|
| **Infection Source** | **n (%)** |
| Bloodstream | 650 (55.2) |
| Urinary tract | 343 (29.1) |
| Respiratory tract | 172 (14.6) |
| Wound | 6 (0.5) |
| Others | 7 (0.6) |
| **City/Hospital** | **n (%)** |
| Riyadh | 593 (50.3) |
| Al-Ahsa | 190 (16.1) |
| Dammam | 86 (7.3) |
| Madinah | 167 (14.2) |
| Jeddah | 142 (12.1) |
| **Collection Year** | **n (%)** |
| 2018 | 240 (20.4) |
| 2019 | 316 (26.8) |
| 2020 | 103 (8.7) |
| 2021 | 111 (9.4) |
| 2022 | 79 (6.7) |
| 2023 | 137 (11.6) |
| 2024 | 192 (16.3) |
| **KPSC Distribution** | **n (%)** |
| *K. pneumoniae* | 1,115 (94.7) |
| *K. quasipneumoniae* | 42 (3.6) |
| *K. variicola* | 11 (0.9) |
| *K. michiganensis* | 4 (0.3) |
| *K. aerogenes* | 4 (0.3) |
| *K. pasteurii* | 2 (0.2) |

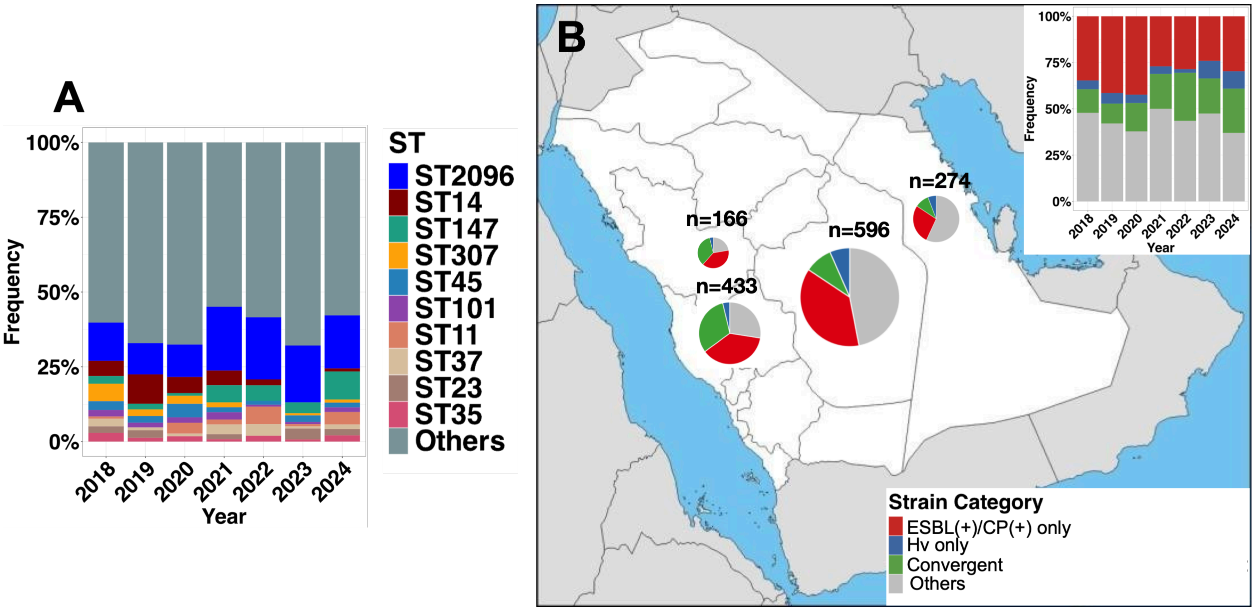


**Figure S1 Distribution of *K. pneumnaie* complex isolates based on A) ST and B ) their risk caterogry (pathotypes) for resistance and virulence across hospitals and years.** We showed STs, represented by more than ten isolates in the collection. The hypervirulent and ESBL/CP states are based on the Kleborate scheme.

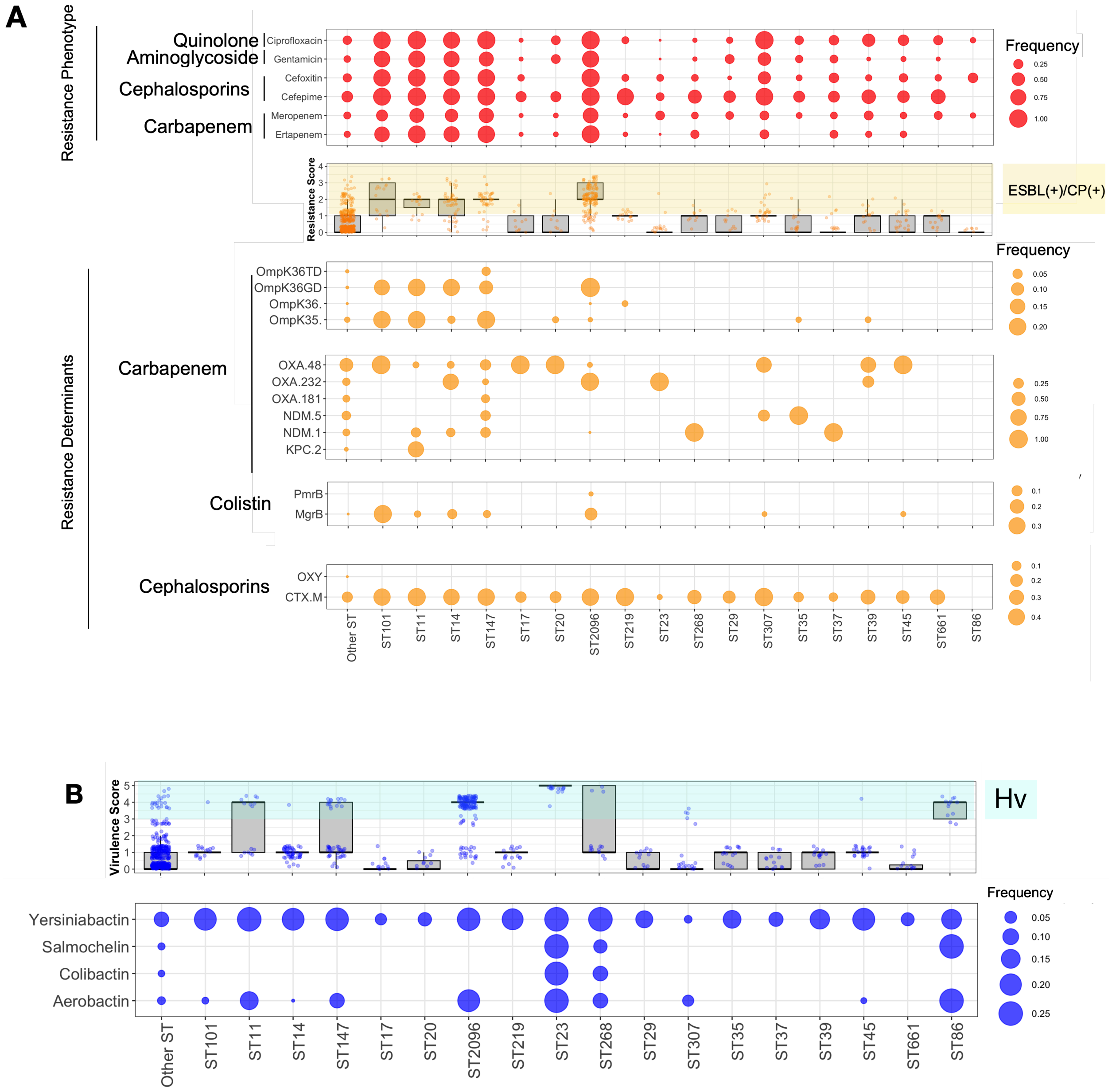


**Figure S2.** The antibiogram, resistome, and virulome profiles across the most frequent clones, i.e., sequence types (STs) represented by at least ten isolates. (A) Proportion of resistant isolates based on antimicrobial susceptibility testing, distribution of resistance scores determined by the Kleborate scoring scheme, and frequency of key resistance genes and mutations conferring resistance to carbapenems, colistin, and cephalosporins. For colistin, the aggregated frequency of mutations in *mgrB* and *pmrB* is shown, while OmpK36 and OmpK35 refer to gene truncations or absence of the OmpK porin genes. The shaded area represents resistance scores $\geq 1$, corresponding to ESBL(+)/CP(+) status. (B) Distribution of virulence scores based on the presence of major hypervirulent genes, along with their relative frequencies across predominant STs. The shaded area indicates hypervirulent status as defined by the Kleborate scoring scheme.

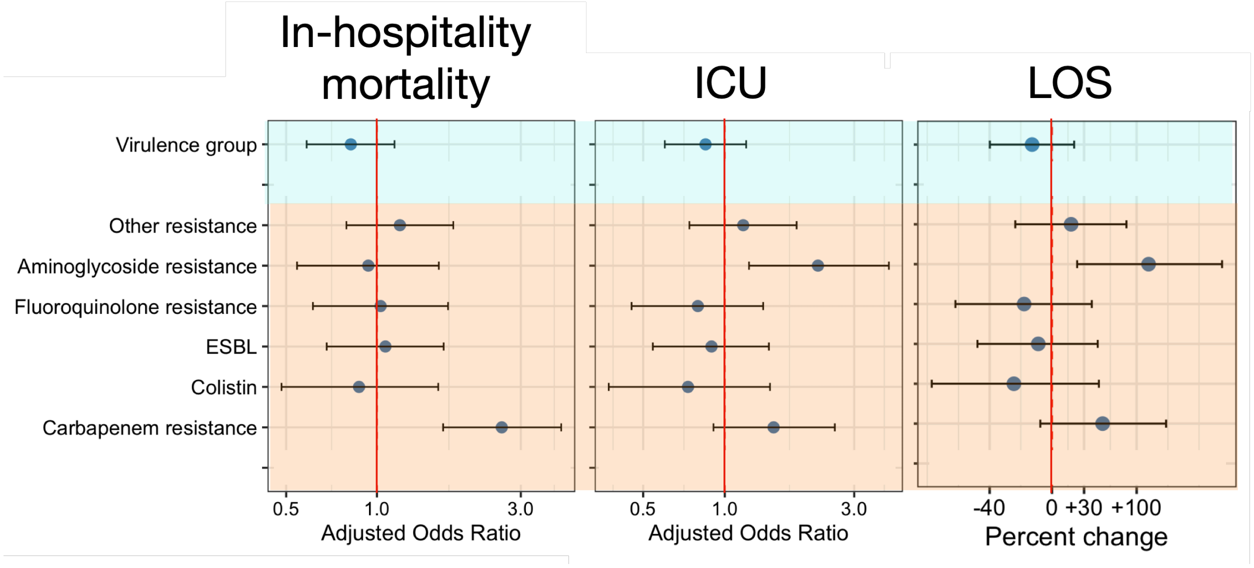


**Figure S3 The association of the virulence and resistance genes with clinical outcome**. We adopted Model 2, in which all genes are incorporated into the multivariable regression model, along with clinical patient-level data so that the effect of each gene can be estimated while controlling for the influence of other resistance or virulence genetic and clinical confounders. The bars denote 95% confidence interval. Genes and mutations are aggregated based on their resistance mechanism. Other resistance refer to all resistance determinants, except ESBLs, colistin and carbapenam resistance determinants.

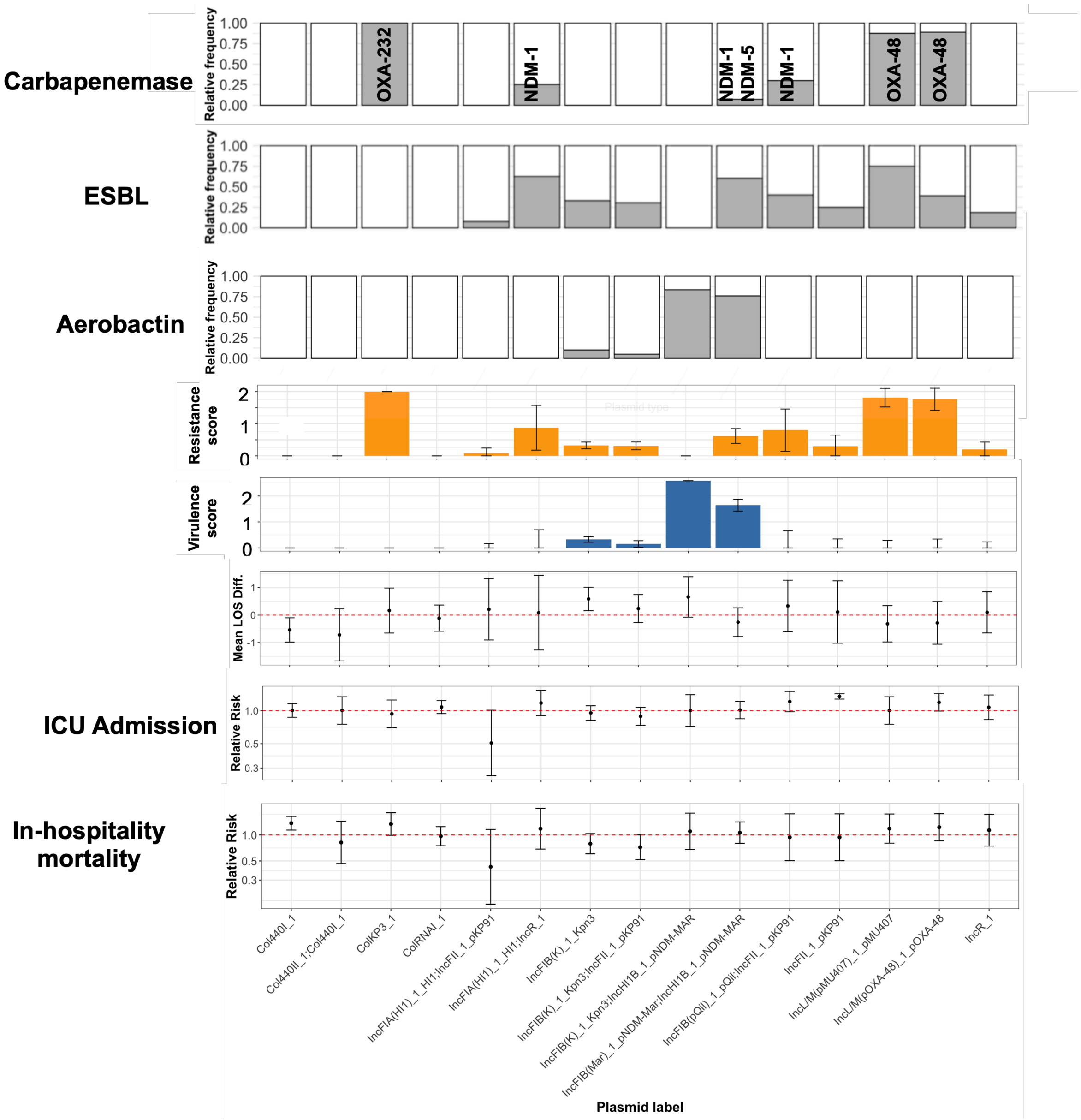


**Figure S4 Risk assessment of plasmid backbone carriage with the clinical outcomes.** The x-axis shows plasmid replicon types identified using the abricate pipeline across 179 extracted plasmid fragments. The upper panels display the relative frequencies of key plasmid-borne resistance and virulence determinants, including carbapenemase genes (*bla*$_{\text{OXA-48}}$, *bla*$_{\text{OXA-232}}$, *bla*$_{\text{NDM-1}}$, *bla*$_{\text{NDM-5}}$) and the aerobactin locus (*iuc*). Middle panels show the mean resistance and virulence scores for each plasmid type, with error bars indicating 95% confidence intervals. The lower panels show the mean difference in length of hospital stay (LOS) and relative risks (95% confidence intervals) for ICU admission and in-hospital mortality. Relative risks were computed using a model wehere only plasmid-containing isolates and their clinical features (confounders) were included.

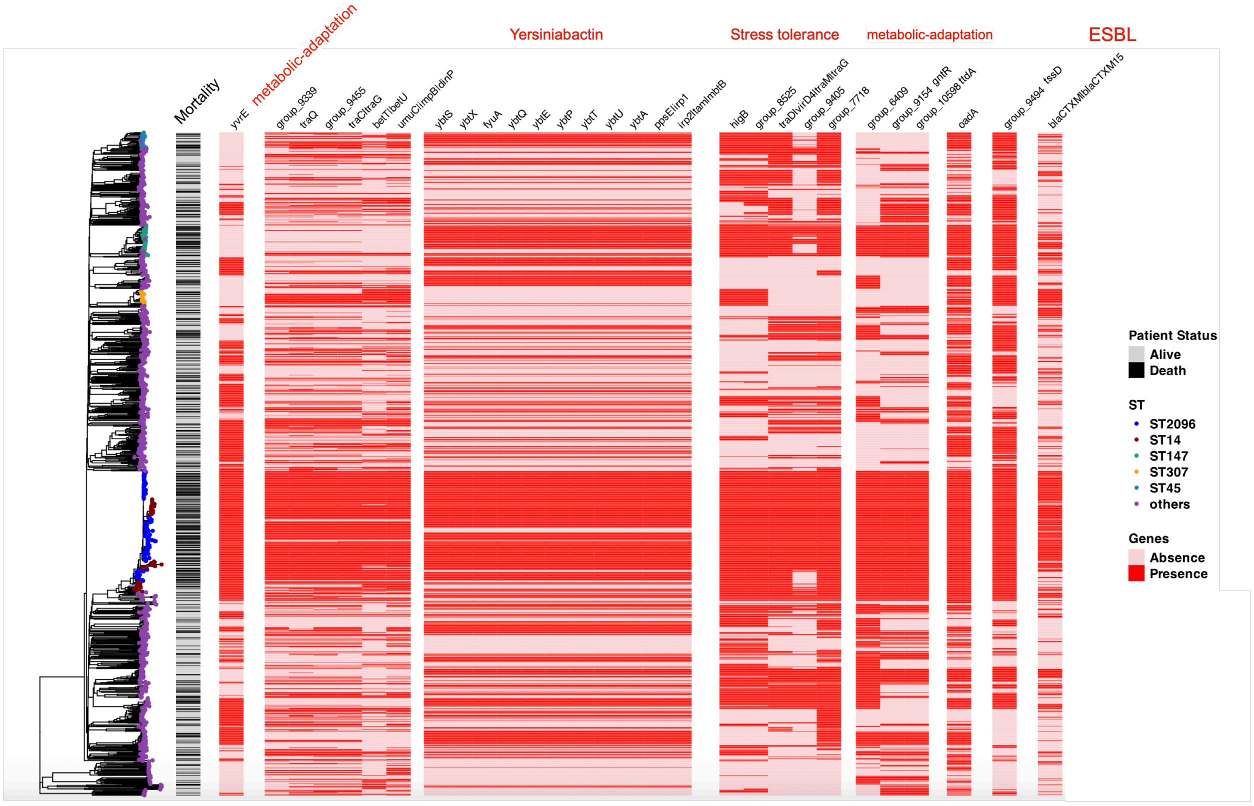


**Figure S5** Phylogenetic distribution of accessory genes significantly linked with in-hospital mortality status. The tree is a neighbor-joining tree for the entire collection reconstructed from pairwise Jaccard kmer distances computed for the entire collection. Accessory gene families were clustered using k-means clustering method, based on their co-occurrence patterns. The annotations and seuqneces of the gene families are provided in Table S3 and the GitHub directory of the study, respectively.

**Table S3** Accessory genes that were significantly associated with clinical outcomes, identified via genome-wide association and regression analyses adjusted for confounders. Sequences for the gene families are provided in the GitHub directory of the project. Rows highlighted in red indicate genes that affect more than one outcome trait.

| Gene | Annotation | Sens | Spec | OR | Trait |
|---|---|---|---|---|---|
| *group_1444* | Gp11 | 37 | 80 | 2.38 | ICU |
| *group_13445* | immunoglobulin domain-containing protein;Ig-like domain-containing protein | 31 | 83 | 2.25 | ICU |
| *group_190* | GNAT family N-acetyltransferase;N-acetyltransferase domain-containing protein | 9 | 97 | 3.66 | ICU |
| *catA1* | Chloramphenicol acetyltransferase;type A-1 chloramphenicol O-acetyltransferase | 9 | 97 | 3.77 | ICU |
| *group_1497* | Helix-turn-helix domain-containing protein;hypothetical protein;helix-turn-helix domain-containing protein | 28 | 85 | 2.24 | ICU |
| *group_3615* | Ead/Ea22-like family protein | 29 | 88 | 2.94 | ICU |
| *group_5476* | DUF6012 domain-containing protein | 29 | 88 | 2.94 | ICU |
| *group_4700* | DUF4165 domain-containing protein;Ig-like domain repeat protein;Ig-like domain-containing protein | 28 | 88 | 2.88 | ICU |
| *group_3674* | Amino acid permease | 29 | 87 | 2.82 | ICU |
| *group_2899* | DUF4942 domain-containing protein | 29 | 88 | 2.87 | ICU |
| *group_9150* | Transmembrane protein;hypothetical protein | 28 | 88 | 2.9 | ICU |
| *group_8148* | Transmembrane protein;hypothetical protein | 29 | 87 | 2.8 | ICU |
| *glpP* | Glycerol-3-phosphate responsive antiterminator (mRNA-binding);Glycerol uptake operon antiterminator regulatory protein | 70 | 42 | 1.68 | LOS |
| *fbaB* | Fructose-bisphosphate aldolase class Ia DhnA family | 70 | 42 | 1.66 | LOS |
| *fabG(group_5655)* | NAD(P)-dependent dehydrogenase short-chain alcohol dehydrogenase family | 70 | 41 | 1.67 | LOS |
| *sgcC(group_12879)* | Phosphotransferase system galactitol-specific IIC component; PTS transporter subunit IIC; PTS system galactitol-specific IIC component;PTS system protein | 70 | 41 | 1.67 | LOS |
| *serA* | Phosphoglycerate dehydrogenase or related dehydrogenase;D-isomer specific 2-hydroxyacid dehydrogenase catalytic domain-containing protein; D-3-phosphoglycerate dehydrogenase | 70 | 41 | 1.65 | LOS |
| *gstA* | Glutathione S-transferase ;glutathione binding-like protein;glutathione S-transferase family protein; Glutathione S-transferase family protein | 82 | 26 | 1.6 | LOS |
| *group_6939* | GIY-YIG nuclease family protein;hypothetical protein | 45 | 65 | 1.5 | LOS |
| *araH(group_6835)* | Ribose/xylose/arabinose/galactoside ABC-type transport system permease component | 53 | 59 | 1.61 | LOS |
| *rbsA\|ccmA* | Ribose import ATP-binding protein RbsA; ATP-binding cassette domain-containing protein; heme ABC exporter ATP-binding protein CcmA | 53 | 59 | 1.61 | LOS |
| *tktA2(group_4927)* | Transketolase C-terminal subunit | 53 | 59 | 1.61 | LOS |
| *tktA1(group_2442)* | Transketolase N-terminal subunit | 53 | 59 | 1.61 | LOS |
| *deoR* | DNA-binding transcriptional regulator LsrR DeoR family | 53 | 59 | 1.61 | LOS |
| *hcp\|tssD(group_9494)* | Type VI protein secretion system component Hcp (secreted cytotoxin); type VI secretion system tube protein TssD | 74 | 39 | 1.82 | LOS |
| *gamL(group_11193)* | host nuclease inhibitor GamL | 51 | 65 | 1.89 | LOS |

| Gene | Annotation | Sens | Spec | OR | Trait |
|---|---|---|---|---|---|
| *virB11(group_760)* | P-type DNA transfer ATPase VirB11 | 53 | 66 | 2.22 | LOS |
| *rbsB(group_3812)* | ABC-type sugar transport system periplasmic component contains N-terminal xre family HTH domain | 53 | 58 | 1.6 | LOS |
| *group_5093* | DUF2857 domain-containing protein | 56 | 55 | 1.6 | LOS |
| *alpA(group_602)* | DNA-binding transcriptional regulator AlpA;AlpA family transcriptional regulator | 56 | 55 | 1.6 | LOS |
| *recE* | RecE family exodeoxyribonuclease;Exodeoxyribonuclease VIII;hypothetical protein; DNA breaking-rejoining protein ;Exodeoxyribonuclease 8;PD-(D/E)XK nuclease-like domain-containing protein; Putative exodeoxyribonuclease VIII;Exodeoxyribonuclease | 57 | 53 | 1.48 | LOS |
| *xerD\|intI1* | Site-specific recombinase XerD;integron integrase;Integron integrase; class 1 integron integrase IntI1 | 58 | 60 | 2.14 | LOS |
| *sul1* | sulfonamide-resistant dihydropteroate synthase Sul1;dihydropteroate synthase | 48 | 71 | 2.18 | LOS |
| *yebW* | Secreted protein;YebW family protein;DUF1482 domain-containing protein | 56 | 54 | 1.49 | LOS |
| *ppsE\|irp1* | Iron aquisition yersiniabactin synthesis enzyme (Irp1polyketide synthetase); yersiniabactin polyketide synthase HMWP1;Non-ribosomal peptide synthase (Yersiniabact siderophore biosynthetic protein); Irp1;Phthiocerol/phenolphthiocerol synthesis polyketide synthase type I PpsE;Carrier domain-containing protein;yersiniabactin biosynthetic protein Irp1 | 57 | 54 | 1.59 | LOS |
| *irp2\|tam\|mbtB* | Amino acid adenylation domain-containing protein;non-ribosomal peptide synthetase; Irp2;Peptide synthetase;Carrier domain-containing protein;AMP-binding enzyme; hypothetical protein; Peptide synthetase-like protein; yersiniabactin non-ribosomal peptide synthetase HMWP2; Trans-aconitate methyltransferase;Phenyloxazoline synthase MbtB | 57 | 54 | 1.59 | LOS |
| *ybtA* | yersiniabactin transcriptional regulator YbtA | 57 | 54 | 1.59 | LOS |
| *ybtU* | Oxidoreductase;yersiniabactin biosynthetic protein YbtU | 57 | 54 | 1.59 | LOS |
| *ybtT* | Thioesterase;Surfactin synthase thioesterase subunit; yersiniabactin biosynthesis thioesterase YbtT | 57 | 54 | 1.59 | LOS |
| *ybtP* | yersiniabactin ABC transporter ATP-binding/permease protein YbtP | 57 | 54 | 1.59 | LOS |
| *ybtE* | yersiniabactin biosynthesis salycil-AMP ligase YbtE; Iron aquisition 23-dihydroxybenzoate-AMP ligase | 57 | 54 | 1.59 | LOS |
| *ybtQ* | yersiniabactin ABC transporter ATP-binding/permease protein YbtQ; Putative ABC transporter protein | 57 | 54 | 1.59 | LOS |
| *fyuA* | yersiniabactin receptor FyuA | 57 | 54 | 1.59 | LOS |
| *ybtX* | Signal transducer;putative signal transducer; yersiniabactin-associated zinc MFS transporter YbtX;MFS transporter | 57 | 54 | 1.59 | LOS |
| *ybtS* | salicylate synthase Irp9;yersiniabactin biosynthesis salicylate synthase YbtS | 57 | 54 | 1.59 | LOS |
| *parB(group_1659)* | PRTRC system ParB family protein;DUF4752 domain-containing protein;hypothetical protein;ParB/Spo0J HTH domain-containing protein;Nucleoid occlusion protein; ParB/RepB/Spo0J family partition protein;ParB/Sulfiredoxin domain-containing protein | 51 | 66 | 2.03 | LOS |
| *virB2(group_1138)* | Major pilus subunit of type IV secretion complex VirB2;TrbC/VirB2 family protein | 55 | 61 | 1.93 | LOS |
| *arr\|arr2* | NAD(+)--rifampin ADP-ribosyltransferase;NAD(+)--rifampin ADP-ribosyltransferase Arr-2 | 11 | 98 | 5.62 | LOS |
| *virB1* | Type IV secretion system protein virB1 | 55 | 61 | 1.91 | LOS |
| *group_9156* | Cold-shock protein | 89 | 18 | 1.81 | LOS |
| *dinI* | DinI family protein;DNA-damage-inducible protein I; XRE family transcriptional regulator; DinI-like family protein;DNA damage-inducible protein I | 49 | 66 | 1.83 | LOS |
| *group_9405* | TIGR03747 family integrating conjugative element membrane protein;Integrating conjugative element membrane protein PFL_4697 family; | 49 | 66 | 1.9 | Mortality |

| Gene | Annotation | Sens | Spec | OR | Trait |
|---|---|---|---|---|---|
| | DUF4400 domain-containing protein | | | | |
| *traC\|traG* | type IV secretion system protein TraC;hypothetical protein;IncF plasmid conjugative transfer p assembly protein TraC;Type IV secretion system protein TraC; TraG P-loop domain-containing protein | 57 | 55 | 1.57 | Mortality |
| *araH(group_6835)* | Ribose/xylose/arabinose/galactoside ABC-type transport system permease component | 55 | 59 | 1.74 | Mortality |
| *rbsA\|ccmA* | Ribose import ATP-binding protein RbsA;ATP-binding cassette domain-containing protein; heme ABC exporter ATP-binding protein CcmA | 55 | 59 | 1.74 | Mortality |
| *tktA2(group_4927)* | Transketolase C-terminal subunit | 55 | 59 | 1.74 | Mortality |
| *tktA1(group_2442)* | Transketolase N-terminal subunit | 55 | 59 | 1.74 | Mortality |
| *deoR* | DNA-binding transcriptional regulator LsrR DeoR family | 55 | 59 | 1.74 | Mortality |
| *higA(group_8525)* | Antitoxin component HigA of the HigAB toxin-antitoxin module contains an N-terminal HTH domain | 66 | 48 | 1.79 | Mortality |
| *yfiB(group_9455)* | YfiB protein | 58 | 54 | 1.6 | Mortality |
| *rbsB(group_3812)* | ABC-type sugar transport system periplasmic component contains N-terminal xre family HTH domain | 55 | 59 | 1.73 | Mortality |
| *higB* | mRNA-degrading endonuclease (mRNA interferase) HigB toxic component of the HigAB toxin-antitoxin module | 65 | 50 | 1.82 | Mortality |
| *group_7718* | DUF2645 domain-containing protein | 72 | 40 | 1.74 | Mortality |
| *group_1561* | Integron gene cassette protein;hypothetical protein | 55 | 58 | 1.69 | Mortality |
| *ttdB* | L(+)-tartrate dehydratase subunit beta;fumarate hydratase C-terminal domain-containing protein | 59 | 59 | 2.02 | Mortality |
| *ttdA(group_10598 )* | L(+)-tartrate dehydratase subunit alpha;hypothetical protein | 59 | 59 | 2.02 | Mortality |
| *citT(group_10069)* | Di- and tricarboxylate antiporter;putative membrane transport protein; Putative cation transporter | 59 | 59 | 2.02 | Mortality |
| *gntR(group_9154)* | DNA-binding transcriptional regulator GntR family | 59 | 59 | 2.02 | Mortality |
| *gntR(group_7317)* | DNA-binding transcriptional regulator GntR family;GntR family transcriptional regulator | 59 | 59 | 2.02 | Mortality |
| *hcp\|tssD(group_9 494)* | Type VI protein secretion system component Hcp (secreted cytotoxin); type VI secretion system tube protein TssD | 74 | 38 | 1.81 | Mortality |
| *ppsE\|irp1* | Iron aquisition yersiniabactin synthesis enzyme (Irp1polyketide synthetase); yersiniabactin polyketide synthase HMWP1;Non-ribosomal peptide synthase (Yersiniabactin siderophore biosynthetic protein);Irp1;Phthiocerol/phenolphthiocerol synthesis polyketide synthase type I PpsE; Carrier domain-containing protein;yersiniabactin biosynthetic protein Irp1 | 59 | 54 | 1.73 | Mortality |
| *irp2\|tam\|mbtB* | Amino acid adenylation domain-containing protein;non-ribosomal peptide synthetase; Irp2; Peptide synthetase;Carrier domain-containing protein; AMP-binding enzyme;hypothetical protein; Peptide synthetase-like protein;yersiniabactin non-ribosomal peptide synthetase HMWP2; Trans-aconitate methyltransferase;Phenyloxazoline synthase MbtB | 59 | 54 | 1.73 | Mortality |
| *ybtA* | yersiniabactin transcriptional regulator YbtA | 59 | 54 | 1.73 | Mortality |
| *ybtU* | Oxidoreductase;yersiniabactin biosynthetic protein YbtU | 59 | 54 | 1.73 | Mortality |
| *ybtT* | Thioesterase;Surfactin synthase thioesterase subunit;yersiniabactin biosynthesis thioesterase YbtT | 59 | 54 | 1.73 | Mortality |
| *ybtP* | yersiniabactin ABC transporter ATP-binding/permease protein YbtP | 59 | 54 | 1.73 | Mortality |
| *ybtE* | ersiniabactin biosynthesis salycil-AMP ligase YbtE;Iron aquisition 23-dihydroxybenzoate-AMP | 59 | 54 | 1.73 | Mortality |

| Gene | Annotation | Sens | Spec | OR | Trait |
|---|---|---|---|---|---|
| *ybtQ* | yersiniabactin ABC transporter ATP-binding/permease protein YbtQ; Putative ABC transporter protein | 59 | 54 | 1.73 | Mortality |
| *fyuA* | yersiniabactin receptor FyuA | 59 | 54 | 1.73 | Mortality |
| *ybtX* | gnal transducer;putative signal transducer;yersiniabactin-associated zinc MFS transporter Ybt transporter | 59 | 54 | 1.73 | Mortality |
| *ybtS* | salicylate synthase Irp9;yersiniabactin biosynthesis salicylate synthase YbtS | 59 | 54 | 1.73 | Mortality |
| *traQ* | type-F conjugative transfer system pilin chaperone TraQ | 58 | 53 | 1.53 | Mortality |
| *oadA* | sodium-extruding oxaloacetate decarboxylase subunit alpha;pyruvate carboxylase subunit B;biotin/lipoyl-containing protein;Oxaloacetate decarboxylase; Pyruvate carboxyltransferase domain-containing protein | 78 | 33 | 1.69 | Mortality |
| *traN(group_9339)* | type-F conjugative transfer system mating-pair stabilization protein TraN; hypothetical protein;Type-F conjugative transfer system mating-pair stabilization protein TraN;conjugal transfer protein TraN;Conjugal transfer mating pair stabilization protein TraN | 54 | 58 | 1.62 | Mortality |
| *citG(group_9545)* | 2-(5''-triphosphoribosyl)-3'-dephosphocoenzyme-A synthase;hypothetical protein; Triphosphoribosyl-dephospho-CoA synthetase;Membrane protein associated with oxaloacetate decarboxylase;oxaloacetate decarboxylase (Na(+) extruding) | 64 | 51 | 1.86 | Mortality |
| *dsbA(group_6409)* | DsbA family protein;Thiol:disulfide interchange protein | 63 | 52 | 1.86 | Mortality |
| *group_498* | DUF905 domain-containing protein;Cytoplasmic protein;hypothetical protein | 53 | 61 | 1.76 | Mortality |
| *umuC\|impB\|dinP* | DNA-directed DNA polymerase;UmuC domain-containing protein; DNA polymerase V protein ImpB;Nucleotidyltransferase/DNA polymerase DinP involved in DNA repair;Error-prone lesion bypass DNA polymerase V (UmuC); Translesion error-prone DNA polymerase V subunit UmuC; translesion error-prone DNA polymerase V subunit UmuC;DNA polymerase V subunit UmuC;hypothetical protein | 57 | 53 | 1.52 | Mortality |
| *ssb(group_472)* | Single-stranded DNA-binding protein;single-stranded DNA-binding protein | 54 | 60 | 1.72 | Mortality |
| *betT\|betU* | CCT family transporter;Choline/carnitine/betaine transporter family protein;Choline-glycine b transporter;Secondary glycine betaine transporter BetU | 49 | 65 | 1.79 | Mortality |
| *group_5625* | Transcriptional regulator;hypothetical protein | 72 | 40 | 1.76 | Mortality |
| *yvrE* | Sugar lactone lactonase YvrE;Gluconolactonase | 62 | 48 | 1.52 | Mortality |
| *blaCTXM\|blaCTX M15* | TX-M family extended-spectrum class A beta-lactamase;extended-spectrum class A beta-lact CTX-M-15 | 51 | 65 | 1.89 | Mortality |
| *group_7720* | Membrane associated protein;hypothetical protein | 49 | 68 | 2.04 | Mortality |
| *group_14243* | Nitrite transporter;nitrite transporter | 29 | 84 | 2.12 | Mortality |

**Table S4.** List of missense and stop-gain SNPs significantly associated with clinical outcomes, identified via genome-wide association and regression analyses adjusted for confounders. The reference genome is BCL8 strain (see Methods). Rows highlighted in red indicate SNPs that affect more than one outcome trait.

| SNPs | Sens | Speci | OR. | LOCUS_TAG | GENE | PRODUCT | Functional Group | Trait |
|---|---|---|---|---|---|---|---|---|
| S92P | 21 | 90 | 2.6 | BN373_09971 | | replication protein | Replication & DNA maintenance | ICU |
| G27D | 24 | 87 | 2.3 | BN373_14121 | | glycosyl hydrolase, family 1 | Metabolism – Carbohydrate degradation | ICU |
| D300G | 27 | 89 | 3 | BN373_31461 | *dalD* | D-arabinitol 4-dehydrogenase | Metabolism – Carbohydrate oxidation | ICU |
| P10S | 40 | 73 | 1.9 | BN373_00681 | | ABC transporter, permease protein | Transport – ABC transporter (permease) | LOS |
| A104D | 3 | 99 | 5.5 | BN373_01711 | *rsgA* | ribosome small subunit-dependent GTPase A | Translation / Ribosome function | LOS |
| D156G | 53 | 64 | 2 | BN373_02391 | | acetyltransferase, GNAT family | Post-translational modification / regulation | LOS |
| I32V | 41 | 70 | 1.6 | BN373_03611 | *cusA* | cation efflux system protein CusA | Transport – Efflux system / metal resistance | LOS |
| R201Q | 24 | 91 | 3.2 | BN373_03781 | | ribose high-affinity ABC transport system permease component | Transport – ABC transporter (substrate uptake) | LOS |
| R271C | 62 | 48 | 1.6 | BN373_04231 | | 2-oxo-3-deoxygalactonate kinase | Metabolism – Carbohydrate metabolism | LOS |
| G356D | 40 | 72 | 1.7 | BN373_10681 | | oxidoreductase, NAD binding | Metabolism – Redox enzyme | LOS |
| I446N | 20 | 87 | 1.8 | BN373_11361 | | oligopeptide/dipeptide ABC transporter, periplasmic oligopeptide/dipeptide-binding protein | Transport – ABC transporter (peptide uptake) | LOS |
| E189K | 48 | 64 | 1.6 | BN373_11991 | | alcohol dehydrogenase, iron-containing | Metabolism – Alcohol/oxidation | LOS |
| F187I | 49 | 63 | 1.7 | BN373_12351 | *pagP* | antimicrobial peptide resistance and lipid A acylation protein PagP | Cell envelope / Resistance & modification | LOS |
| R66H | 43 | 67 | 1.6 | BN373_14341 | *bssR* | biofilm regulator BssR | Regulation – Biofilm formation | LOS |
| S97C | 59 | 52 | 1.6 | BN373_15991 | *sulA* | cell division inhibitor SulA | Cell cycle / Division control | LOS |
| R37* | 4 | 99 | 5.2 | BN373_18711 | *ompW* | outer membrane protein W | Cell envelope – Outer membrane protein | LOS |
| A40T | 41 | 69 | 1.6 | BN373_19481 | | intracellular protease, PfpI family | Protein turnover / Protease | LOS |
| D37E | 3 | 99 | 7.7 | BN373_19791 | | NmrA family protein | Regulation – Sensor/regulator | LOS |
| K535E | 73 | 36 | 1.6 | BN373_21011 | | transporter, small conductance mechanosensitive ion channel (MscS) family | Transport – Ion channel / mechanosensitive | LOS |
| I262V | 57 | 55 | 1.7 | BN373_22391 | | transporter, major facilitator family | Transport – Major facilitator superfamily (MFS) | LOS |
| A93T | 3 | 99 | 6.6 | BN373_22741 | | major facilitator family transporter | Transport – Major facilitator superfamily (MFS) | LOS |

| SNPs | Sens | Speci | OR. | LOCUS_TAG | GENE | PRODUCT | Functional Group | Trait |
|---|---|---|---|---|---|---|---|---|
| T237K | 63 | 48 | 1.6 | BN373_23561 | | class I glutamine amidotransferase family protein | Metabolism – Amino acid / amide metabolism | LOS |
| A199E | 22 | 88 | 2.1 | BN373_23751 | | TENA/THI-4 family protein | Miscellaneous – Co-factor / unknown function | LOS |
| S248R | 44 | 71 | 1.9 | BN373_25901 | | transcriptional regulator, LysR family | Regulation – Transcriptional regulator (LysR) | LOS |
| K423N | 4 | 99 | 5.2 | BN373_26671 | | gluconate 2-dehydrogenase, cytochrome c subunit | Metabolism – Carbohydrate oxidation / electron transport | LOS |
| I164V | 55 | 56 | 1.6 | BN373_27081 | *mhpR* | Mhp operon transcriptional activator | Regulation – Operon transcription activator | LOS |
| D44E | 57 | 54 | 1.6 | BN373_30181 | | ErfK/YbiS/YcfS/YnhG family protein | Miscellaneous – Uncharacterized / accessory | LOS |
| V123L | 4 | 99 | 5.2 | BN373_30761 | | NAD dependent epimerase/dehydratase family protein | Metabolism – Redox / epimerase / dehydratase | LOS |
| Y31F | 63 | 46 | 1.5 | BN373_31021 | *cpsB_1* | mannose-1-phosphate guanylyltransferase/mannose-6-phosphate isomerase | Metabolism – Sugar nucleotide / isomerase | LOS |
| Q80* | 4 | 99 | 4.4 | BN373_31371 | | ABC transporter, ATP-binding protein | Transport – ABC transporter (ATP binding) | LOS |
| S190N | 55 | 58 | 1.7 | BN373_34901 | | transporter, major facilitator family | Transport – Major facilitator superfamily (MFS) | LOS |
| N1403T | 39 | 73 | 1.8 | BN373_35071 | | alpha-2-macroglobulin domain protein | Defense / Protease inhibitor / stress response | LOS |
| Q400H | 55 | 55 | 1.6 | BN373_37011 | *fhlA* | formate hydrogenlyase transcriptional activator | Regulation – Metabolic transcriptional activator | LOS |
| A86T | 5 | 99 | 3.8 | BN373_37961 | *xni* | exodeoxyribonuclease-9 | DNA repair / maintenance | LOS |
| R173Q | 57 | 53 | 1.6 | BN373_38301 | *cobB* | cobyrinic acid a,c-diamide synthase | Metabolism – Cofactor / Vitamin B12 biosynthesis | LOS |
| I228V | 48 | 62 | 1.5 | BN373_38451 | *pduO* | propanediol utilization protein, PduO | Metabolism – Propanediol utilization / cofactor enzyme | LOS |
| T575S | 4 | 99 | 5.2 | BN373_38581 | *recD* | exodeoxyribonuclease V, alpha subunit | DNA repair / maintenance | LOS |
| S85N | 73 | 36 | 1.6 | BN373_39991 | *scsB* | suppressor for copper-sensitivity B | Resistance / Metal homeostasis | LOS |
| S422N | 62 | 49 | 1.6 | BN373_45781 | *malS* | alpha-amylase, periplasmic | Metabolism – Carbohydrate degradation | LOS |
| P24Q | 44 | 67 | 1.7 | BN373_49231 | | glycosyl transferase, WecB/TagA/CpsF family | Cell envelope – Glycosylation / polysaccharide biosynthesis | LOS |
| H729Q | 60 | 55 | 1.9 | BN373_03611 | *cusA* | cation efflux system protein CusA | Transport – Efflux system / metal resistance | Mortality |
| A180E | 43 | 71 | 1.8 | BN373_11411 | | amidase, hydantoinase/carbamoylase family | Metabolism – Amide metabolism / hydantoinase activity | Mortality |
| E192D | 61 | 55 | 1.9 | BN373_11411 | | amidase, hydantoinase/carbamoylase family | Metabolism – Amide metabolism / hydantoinase activity | Mortality |

| SNPs | Sens | Speci | OR. | LOCUS_TAG | GENE | PRODUCT | Functional Group | Trait |
|---|---|---|---|---|---|---|---|---|
| Q461K | 49 | 65 | 1.9 | BN373_12191 | | sugar ABC transporter, ATP-binding protein | Transport – ABC transporter (sugar uptake) | Mortality |
| V650I | 53 | 64 | 2 | BN373_12211 | | biotin sulfoxide reductase homolog | Metabolism – Redox / cofactor maintenance | Mortality |
| A40T | 44 | 70 | 1.9 | BN373_19481 | | intracellular protease, PfpI family | Protein turnover / Protease | Mortality |
| H56Y | 61 | 53 | 1.8 | BN373_19781 | | metallo-beta-lactamase domain protein | Resistance / Detoxification enzyme | Mortality |
| V98I | 43 | 80 | 3 | BN373_19821 | | allinase | Metabolism – Amine / sulfur compound degradation | Mortality |
| V250I | 59 | 55 | 1.8 | BN373_22391 | | transporter, major facilitator family | Transport – Major facilitator superfamily (MFS) | Mortality |
| R263G | 62 | 53 | 1.9 | BN373_24371 | | ABC transporter, ATP-binding protein | Transport – ABC transporter (ATP binding) | Mortality |
| V55L | 43 | 71 | 1.8 | BN373_26781 | | glyoxalase family protein | Metabolism – Detoxification / glyoxal repair | Mortality |
| I164V | 60 | 58 | 2.1 | BN373_27081 | *mhpR* | Mhp operon transcriptional activator | Regulation – Operon transcriptional activator | Mortality |
| S252A | 66 | 49 | 1.9 | BN373_34521 | *eutT* | cob(I)yrinic acid a,c-diamide adenosyltransferase | Metabolism – Cofactor / Vitamin B12 biosynthesis | Mortality |
| T24A | 38 | 78 | 2.3 | BN373_34621 | *acrD* | acriflavine resistance protein D | Resistance / Drug efflux or modification | Mortality |
| S190N | 57 | 58 | 1.9 | BN373_34901 | | transporter, major facilitator family | Transport – Major facilitator superfamily (MFS) | Mortality |
| S422N | 65 | 50 | 1.9 | BN373_45781 | *malS* | alpha-amylase, periplasmic | Metabolism – Carbohydrate degradation | Mortality |

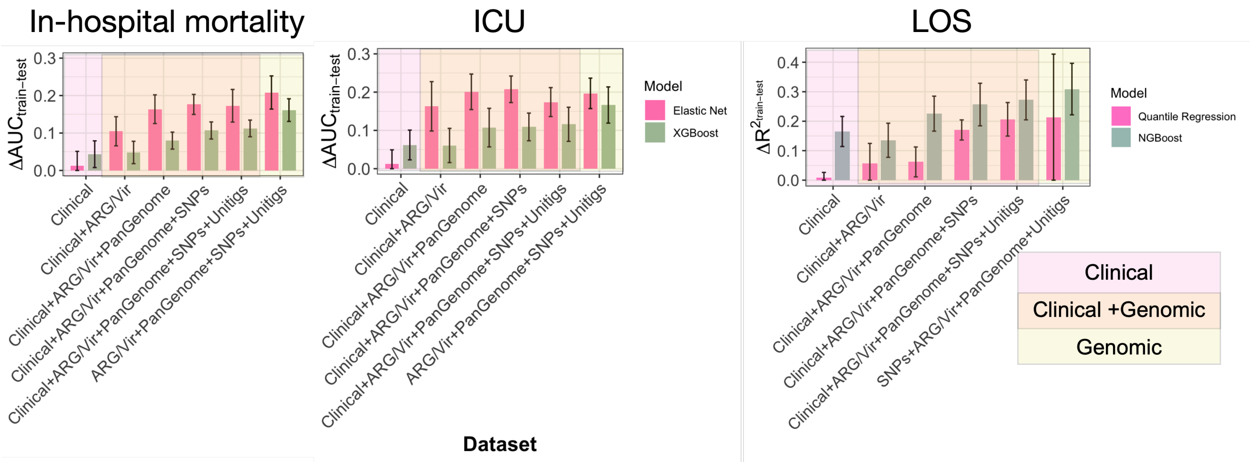


**Figure S6 Training–test performance divergence across feature sets.** A) For in-hospital mortality and ICU admission classification, elastic net (baseline) and XGBoost (ensemble) models were evaluated. The difference in AUC between training and test sets (ΔAUC) was computed across five stratified folds as a measure of training–test performance divergence. B) For LOS prediction, baseline quantile regression and NGBoost models were assessed, and the difference in $R^2$ between training and test sets ($\Delta R^2$) was computed analogously. Error bars represent the standard deviation of ΔAUC (A) or $\Delta R^2$ (B), estimated by propagating fold-wise uncertainties using the root-sum-square method. While increased divergence reflects greater model complexity with the inclusion of genomic features, close agreement between validation and test performance (Figure 7) indicates that this divergence does not correspond to substantial overfitting.

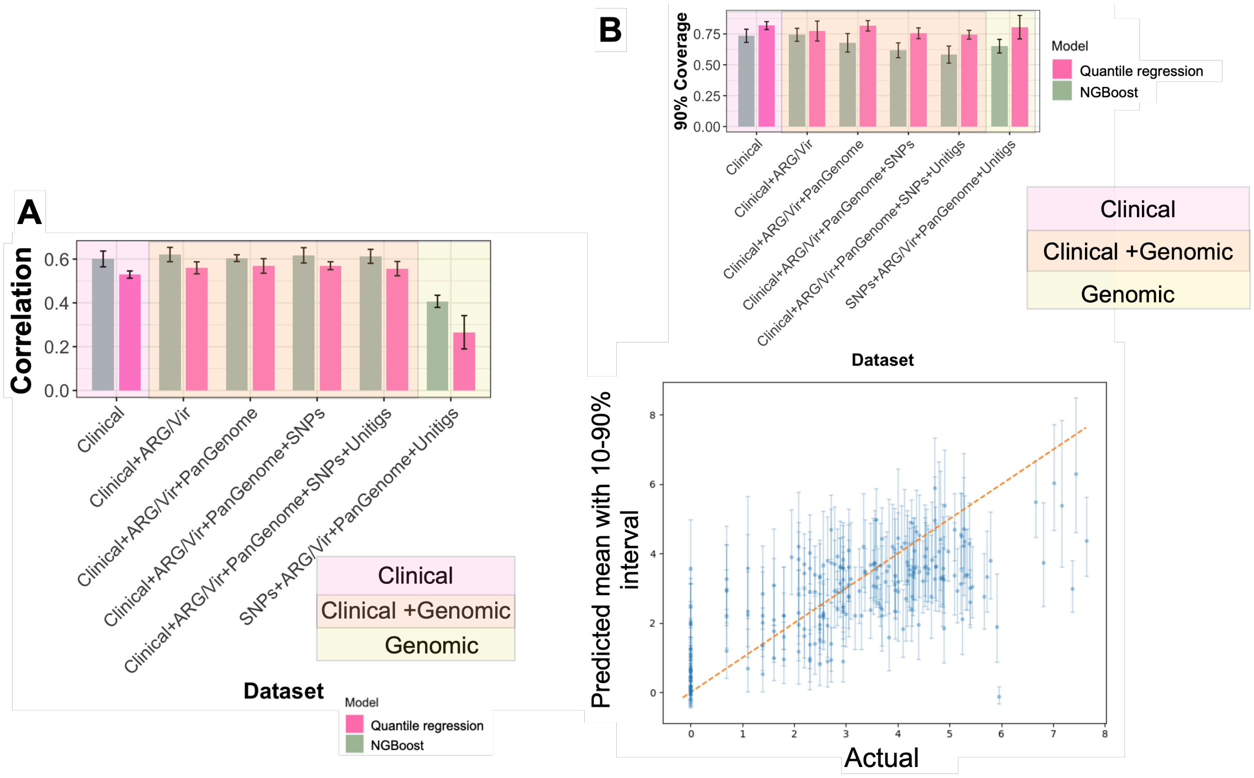


**Figure S7. Point and interval LOS prediction from clinical and genomic data.** A) Model comparison of quantile regression and NGBoost across different feature combinations. The panel shows Pearson's correlation coefficients (*r*) between predicted and observed values. Feature sets include clinical variables, antimicrobial resistance genes (ARGs), virulence genes (Vir), pan-genome data, SNPs, and contig-based (unitig) features. Error bars denote standard deviations across cross-validation folds. B) The upper panel shows interval prediction accuracy measured as the empirical coverage of 90% prediction intervals for input features for quantile regressor and NGBoost models. The bottom panel shows predicted versus actual LOS using the NGBoost model with all genomic feature predictor input data, with the 10–90% predictive interval shown as vertical error bars. The 10–90% predictive interval represents the range within which the model predicts the true value will fall with 80% probability, excluding the most extreme 10% of predictions at each tail. The dashed orange line indicates perfect prediction (y = x).

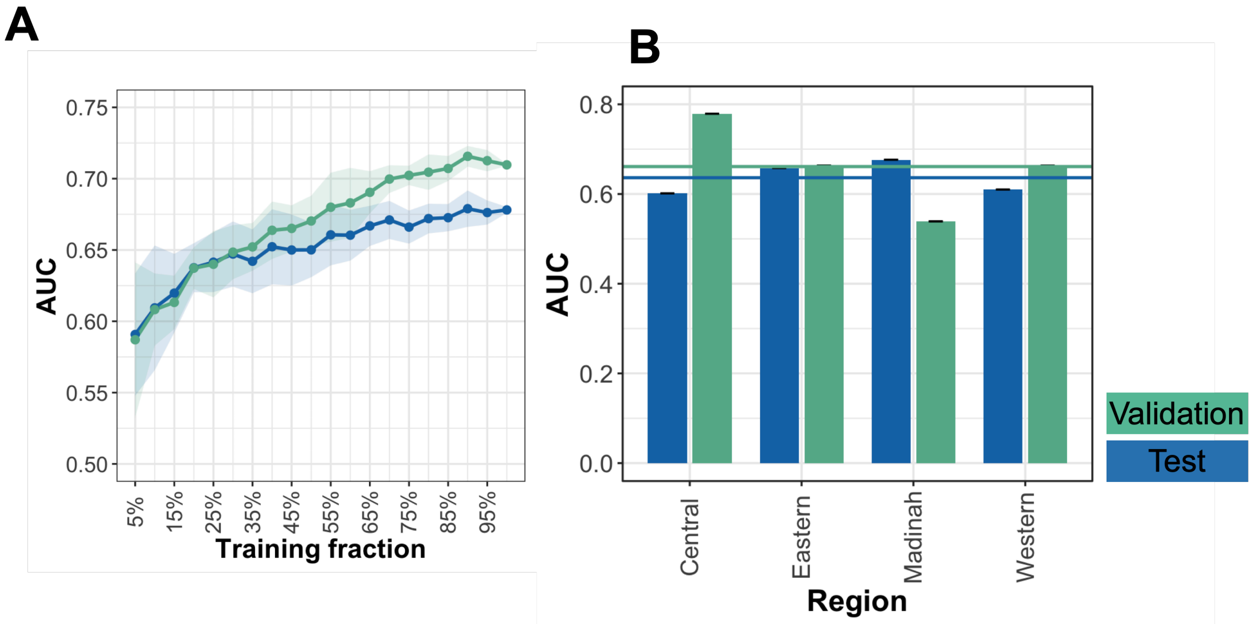


**Figure S8 Model performance analysis. A)** Performance of the XGBoost model evaluated as the AUC values across test and validation subsets derived from different fractions of the training data (x-axis), to assess the impact of training size on model accuracy. The model was trained on combined clinical and all genomic features for ten times on randomly selected subsets. The line shows the mean values across ten runs and the shaded region shows 95% confidence inetrval. **B)** Generalizability of the XGBoost model evaluated across geographic regions. For each run, the model was trained on isolates from all but one region and tested on strains from the held-out region, with training and test sets balanced through random subsampling. Mean AUC values from five cross-validation runs are shown as horizontal lines, and error bars represent the standard deviation across runs.

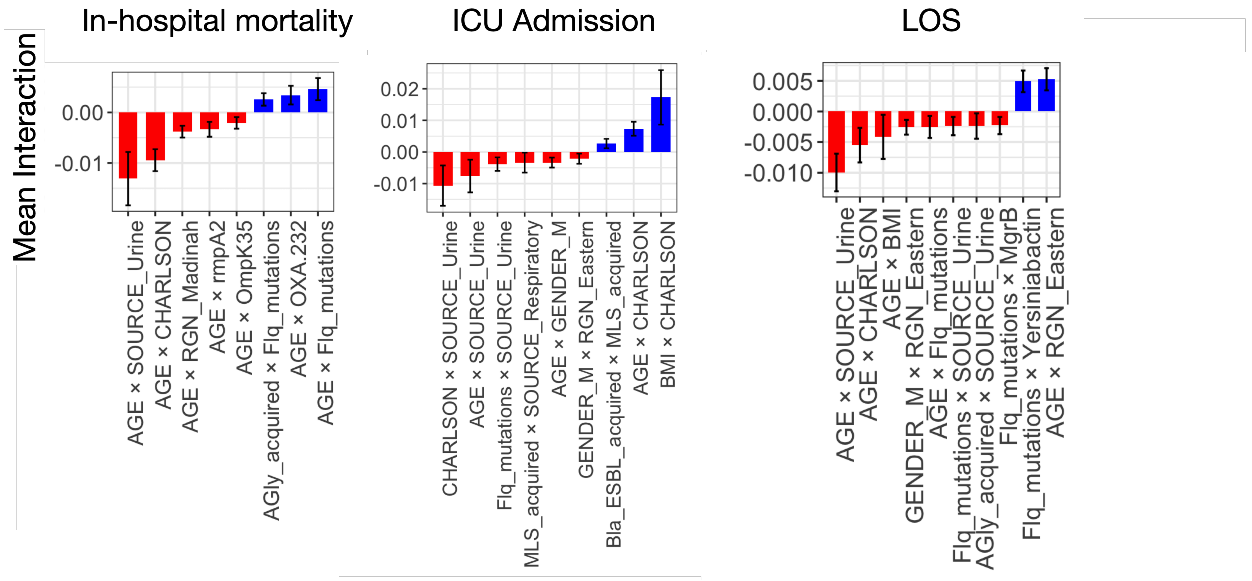


**Figure S9.** Feature interactions for predicting clinical outcomes, i.e. in-hospital mortality, ICU admission status, and LOS, based on clinical features and resistance and virulence gene profiles. Bars represent the mean SHAP interaction values across ten independent runs, and error bars represent 95% confidence inetrvals. Only interactions with significant values, i.e. those whose 95% confidence intervals did not cross zero, are shown. Abbreviations are as follows: BMI, body mass index; CHARLSON, Charlson comorbidity index; SOURCE_Urine and SOURCE_Respiratory, urinary and respiratory infection source; RGN_Eastern and RGN_Madinah, Eastern and Madinah regions; GENDER_M, male sex; FQ_mutations, fluoroquinolone-resistance mutations; AGly_acquired, acquired aminoglycoside-resistance determinants; MLS_acquired, acquired macrolide–lincosamide–streptogramin resistance determinants; bla_ESBL_acquired, acquired ESBL genes; OXA-232, $\textit{bla}_{\text{OXA-232}}$ carbapenemase gene; OmpK35, mutations in the *ompK35* porin gene.
